# Precision medicine for pandemics: stratification of COVID-19 molecular phenotypes defined by topological analysis of global blood gene expression

**DOI:** 10.1101/2024.04.15.24305820

**Authors:** Rebekah Penrice-Randal, Fabio Strazzeri, Benoit Ernst, Brice Van Eeckhout, Julien Guiot, Anna Julie Peired, Cosimo Nardi, Erika Parkinson, Monique Henket, Alicia Staderoli, Elora Guglielmi, Laurie Giltay, Rebecca Baker, Kit Howard, Catherine Hartley, Tessa Prince, Thomas Kleyntssens, Tommaso Manciulli, Ratko Djukanovic, Tristan Clark, Diana Baralle, Scott S Wagers, Xiaodan Xing, Yang Nan, Shiyi Wang, Guang Yang, Paul J Skipp, Julian A Hiscox, James P R Schofield

## Abstract

Precision medicine offers a promising pathway for improving therapeutic responses to pandemics like COVID-19. This study utilises independent patient cohorts from Florence and Liège, collected under the DRAGON consortium, to classify molecular phenotypes associated with COVID-19 through topological analysis of whole blood gene expression. Samples from 173 patients were collected, and RNA was sequenced using the Novaseq platform. Molecular phenotypes were identified through topological analysis of gene expression in relation to the biological network using the TopMD algorithm. Clustering of patients’ topological maps of differential pathway activation uncovered three distinct molecular phenotypes of COVID-19 in the Florence cohort, which were also found in the Liège cohort.

Cluster 1 was characterised by high activation of pathways linked to ESC pluripotency, NRF2, and TGF-β receptor signalling. Cluster 2 showed high activation of pathways, including focal adhesion-PI3K-Akt-mTOR signalling and type I interferon induction and signalling, while Cluster 3 displayed low IRF7-related pathway activation. TopMD was also used alongside the Drug-Gene Interaction Database (DGIdb), revealing pharmaceutical interventions that target mechanisms across multiple phenotypes and individuals.

The data demonstrate the usefulness of molecular phenotyping through topological analysis of blood gene expression and hold promise for guiding personalised therapeutic strategies, not only for COVID-19 but also for Disease X. Its potential for transferability across various diseases emphasises its value in pandemic response efforts, providing insights before large-scale clinical studies are undertaken.

## Introduction

The ongoing challenges of COVID-19, caused by the emergence of SARS-CoV-2, require a thorough understanding of disease heterogeneity. Despite extensive research characterising the host response to SARS-CoV-2 through pre-clinical and clinical ^1,2^ functional genomic data, few approaches have utilised data that cover the full spectrum of symptom severity and disease heterogeneity, and that deliver personalised medicine.

Examination of gene expression patterns in blood has been used in previous studies to identify molecular phenotypes associated with different disease profiles in several emerging viral infections, including Ebola virus (EBOV) ^3^ and SARS-CoV-2 ^1,2,4–6^, as well as more endemic infections such as influenza virus^5^. Medical countermeasures focus on either reducing viral load through anti-virals. These target viral biology or modulate the host response to infection, thereby decreasing sequelae such as inflammation. With any emerging viral pathogen, direct-acting antivirals take time to develop and undergo trials. Identifying therapeutics that can modulate the host response to reduce symptomatology remains a priority. Being able to characterise aberrations in host pathways that lead to disease quickly and integrating this with therapeutics on the FDA-approved list will improve pandemic preparedness and rapid response. Therefore, a deeper understanding of the host response can guide the selection of host-directed medication countermeasures.

The field of digital health and precision medicine is rapidly evolving, with emerging technologies and initiatives aimed at integrating diverse datasets to inform clinical decision-making. In this study, we present a novel approach to analysing complex data collected by the DRAGON international consortium, which enables the rapid identification of targets for treatment by novel and/or repurposed drugs. Within DRAGON, efforts have been made to harmonise data in digital healthcare, to identify biomarkers^7^ and to propose guidelines for the integration of clinical data from various modalities^8^. Additionally, an online platform has been developed to host validated COVID-19 predictive models, facilitating their utilisation by clinicians in real-time decision-making^9^. However, challenges persist, as evidenced by the limited success of outcome prediction models for COVID-19 patients based on demographic and comorbidity data, which highlights the need for more sophisticated approaches^10^.

While omics data have been instrumental in advancing our understanding of SARS-CoV-2 and COVID-19, their integration into digital health platforms for clinical decision-making remains limited^11–13^. Traditional molecular phenotyping approaches often provide only shallow insights. In previous work, using topological analysis, we demonstrated how gene expression data derived from whole blood at the time of admission could predict ICU admission^6^. However, the current study analysed the blood transcriptomes of patients with COVID-19 as part of the DRAGON-EU consortium and used TopMD. This algorithm considers all available data across a landscape of pathways to characterise molecular phenotypes of COVID-19 patients admitted to the hospital. Pathways were identified that correlated with clinical disease in the patient cohort. TopMD mapped pathways onto a database containing information on FDA-approved drugs and their known gene and pathway interactions to generate a list of potential therapeutics for modulating the severity of COVID-19. The ability to rapidly identify and therapeutically modulate host pathways responsible for disease, utilising pre-existing medical countermeasures, will be crucial in responding to the emergence of novel diseases and future pandemics.

This study describes an analysis of the blood transcriptomes of patients with COVID-19 admitted to hospitals in Liège and Florence between February and July 2021, as part of the DRAGON-EU consortium. Alongside collecting blood samples, demographic and clinical observations were recorded; additionally, CT scan data were obtained for a subset of these patients. We applied an unsupervised approach, in which we characterised the molecular phenotypes of patients within this cohort. We have previously reported the development of a gene signature in patients with COVID-19 that predicts admission to the ICU^6^. This predictive signature revealed the activation of pathways regulating epidermal growth factor receptor (EGFR) signalling, peroxisome proliferator-activated receptor alpha (PPAR-α) signalling and transforming growth factor beta (TGF-β) signalling. The observed molecular phenotype aligns with the mechanisms implicated in pulmonary fibrosis, which is also associated with increased severity of disease^14–16^.

## Methods

### Study population and sample collection and ethics

Blood samples were collected between February and July 2021 from 132 COVID-19 patients admitted to Careggi University Hospital in Florence and 41 from a predefined cohort in Liège. All tested positive for SARS-CoV-2 via nasopharyngeal PCR, and samples were drawn on Day 0 of hospital admission. The study was approved by the ethics committees of the University Hospital of Liège (2021/89) and UNIFI (#18085/OSS), with informed consent obtained from all participants.

### Ethical Approval statement

All procedures adhered to the Declaration of Helsinki, relevant laws, and institutional guidelines, with approval from the appropriate ethics committees. Informed consent was obtained from all participants. Clinical data—including age, sex, BMI, and comorbidities— were extracted from electronic medical records and formatted according to the CDISC Study Data Tabulation Model (SDTM).

### Chest CT analysis

Chest CT data were available for 109 of the 173 patients with RNA-seq data. Scans were acquired using a 128-detector multislice Spiral CT (Somatom Definition AS, Siemens Healthcare, Erlangen, Germany) with the following parameters: 150 mAs, 100 kV, 0.3 s rotation, 1.2 mm pitch, 0.465 mm pixel size, 128 × 0.6 mm collimation, 1 mm slice thickness and reconstruction, and Bf70 very sharp kernel. Axial images were captured from the lung apex to base during full inspiration with a breath hold. Coronal and sagittal 1-mm reconstructions were oriented on the tracheal plane. No intravenous contrast was used. Images were reviewed on a 24-inch, 3-megapixel Barco medical monitor (2048 × 1536 resolution). Native MSCT software was used for image analysis, including evaluation of scan quality (inspiratory level and motion artefacts). CT-derived metrics included:

- **CO-RADS score** (1–5): Reflects likelihood of COVID-19 lung involvement (1: very low; 2: low; 3: equivocal; 4: high; 5: very high).
- **Lobar involvement score** (0–5): 0 (0%), 1 (<5%), 2 (5–25%), 3 (26–50%), 4 (51–75%), 5 (>75%).
- **Dominant CT pattern**: Ground-glass opacities, consolidations, mixed, crazy-paving, reverse halo (per Fleischner Society).
- **Distribution**: Lower/upper lobes, peripheral, bronchocentric, dorsal, diffuse.
- **Additional COVID-19-related findings**: Pleural thickening, vascular enlargement, subpleural sign, halo sign, air bubble sign, perilobular pattern, subpleural sparing.
- **Non-typical findings**: Pleural/pericardial effusion, lymphadenopathy, cavitation, tree-in-bud, small nodules, isolated consolidation, atelectasis, septal thickening.

### RNA extraction

Total RNA was extracted from PAXgene Blood RNA Tubes using the PAXgene Blood RNA Kit (PreAnalytix), following the manufacturer’s protocol, and stored at –80°C. RNA was processed using the QIAseq FastSelect–rRNA/Globin Kit (Qiagen) to remove cytoplasmic/mitochondrial rRNA, as well as globin mRNA, with fragmentation times of 7 or 15 minutes. Libraries were prepared using the NEBNext Ultra II Directional RNA Library Prep Kit (New England Biolabs), with either 11 or 13 amplification cycles, followed by AMPure XP bead purification. Libraries were quantified by Qubit, assessed with the Agilent 2100 Bioanalyzer, pooled in equimolar ratios, and sequenced as 150 bp paired-end reads on an Illumina® NovaSeq 6000.

### Bioinformatics

Raw FASTQ files were trimmed using fastp^17^. Trimmed reads were quantified with salmon (v1.5.2) using -l A --validateMappings --seqBias --gcBias^18^. Resulting quantification files were imported into RStudio (v4.1.1) with tximport, and normalised using the edgeR package (v3.34.1)^19^. Sequencing data are available under BioProject ID: PRJNA1085259 on the SRA.

### TopMD molecular phenotype mapping

Topological analysis was performed using TopMD (Patent: GB202306368D0), which maps global gene expression onto a STRING-derived interaction network^20^. Each gene was assigned to a network vertex, and TopMD identified topological features—hotspots of differential expression—representing modulated gene pathways. Pathway activation was quantified as topological volume: the sum of squared differential expression of genes in each cluster. The resulting molecular phenotype is defined as the global profile of pathway activation volumes.

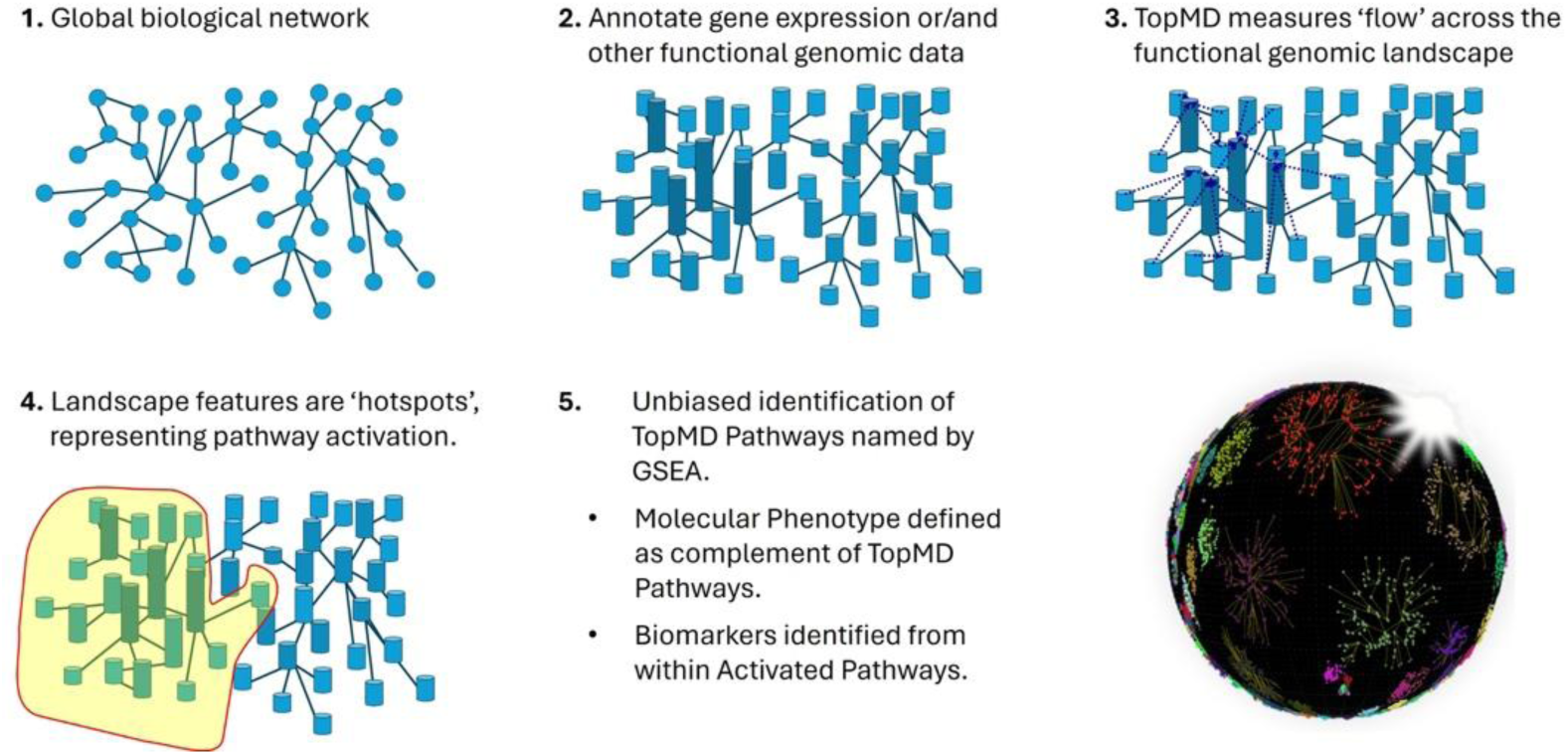

### Drug interactions mapped by topological analysis

TopMD gene clusters were compared to the Drug-Gene Interaction Database (DGIdb)^21^, which catalogues known or predicted drug–gene interactions. Overlap between TopMD gene groups and drug-associated genes was assessed using a binomial distribution test to calculate the probability of random overlap. Results included the raw and Bonferroni-adjusted p-values, TopMD volume (integrating pathway volume with drug-gene overlap significance), and an activation value (sum of Log2 fold-changes for genes shared between the TopMD cluster and the drug-associated set).

### Regression

Regression analysis was performed using a Logistic regression model with the following optimisation problem:

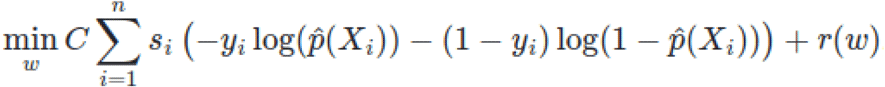

Where X is the **pathway matrix** and y is the classification vector, with 0 indicating that the i-th sample belongs to the considered class and 1 otherwise. We used a regularisation parameter C value of 1. For the penalisation term *r(w)* for the regression weights w, we applied an ElasticNet penalisation with an **l1 ratio** parameter value of 0.5.

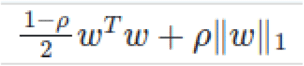

The probability the i-th sample with **pathways values** equal to Xi is then:

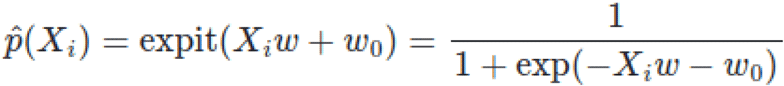

With w0 the intercept. The python module used was **scikit-learn (version 1.4.1)** and the algorithm used LogisticRegression function in the linear_model submodule.

A 70/30 stratified split was carried out separately for each cohort and repeated over 10 random iterations. For each split, regression models were trained using between 1 and 20 pathways, ranked by pathway volume within that split. Each model’s performance was assessed by averaging AUC scores across training and test sets. The best model was selected using a max-min criterion—maximising the minimum AUC between training and test sets—to prevent models that perform poorly in training but seem effective on test data due to random variation.

### Patient Clustering

Pathway volumes were visualised through PCA using PCAtools (v2.14.0), revealing three distinct clusters, which were confirmed by K-means clustering based on pathway activation relative to healthy controls. The top 10% of PCA loadings were extracted to identify pathways responsible for cluster separation. To compare pathway activity between clusters, the average volume of each pathway within each cluster was calculated relative to the overall COVID-19 patient average.

Logistic Regression Receiver Operating Characteristic (LRROC) analysis using patient clusters derived from the patient pathway volume matrix.

The area under the ROC curve (AUC) gauges a model’s capacity to differentiate between classes, with higher AUC scores signifying better discrimination and more clearly defined patient groups.

### LRROC for Florence patients

Logistic regression ROC (LRROC) analysis was conducted using the pathway volume matrix from Florence patients. The dataset was split into training and test sets to assess generalisation, and separate ROC curves were generated for each.

Cluster validation for Liège patients:

The LRROC model trained on Florence data was validated using Liège patient data. An ROC curve was generated to evaluate the model’s ability to classify Liège patient clusters based on pathway volume.

## Results

RNA sequencing was conducted on peripheral blood samples from 173 patients recruited through the DRAGON consortium, with 132 from Florence and 41 from Liège*. The aim was* to evaluate the potential of combining blood transcriptomics with clinical data using the TopMD machine learning approach. Patient details are listed in Supplementary Table 1. Ten patients experienced fatal outcomes; however, due to limited statistical power, an unsupervised analysis was utilised. CT scans, scored by clinicians, were available for 109 patients (Supplementary Table 2). Most had high or very high CORADS scores, while 26% were equivocal, 4.6% low, and 2.8% very low. CORADS (“COVID-19 Reporting and Data System”) is a CT-based classification to estimate the likelihood of COVID-19 infection, ranging from very low to very high.

### Patients form three clusters based on pathway activation

Gene expression (mRNA identity and abundance) was obtained from RNA sequencing using Salmon and Tximport in R, with values converted to log2 counts per million (CPM). Pathway activation was then determined using TopMD. Principal component analysis (PCA) of pathway activation identified three distinct patient clusters (Figure 1). Clinical features, demographics, and CT scan data were compared across these clusters, with significant differences summarised in Table 3. Lactic acid levels were slightly higher in clusters 1 and 2, and lower in cluster 3. Cluster 2 showed a higher prevalence of respiratory disease, increased fraction of inspired oxygen, and elevated direct bilirubin. Most COVID-19-related deaths occurred within cluster 2. CORADS scores did not distinguish the clusters at the molecular level.

**Figure 1:**
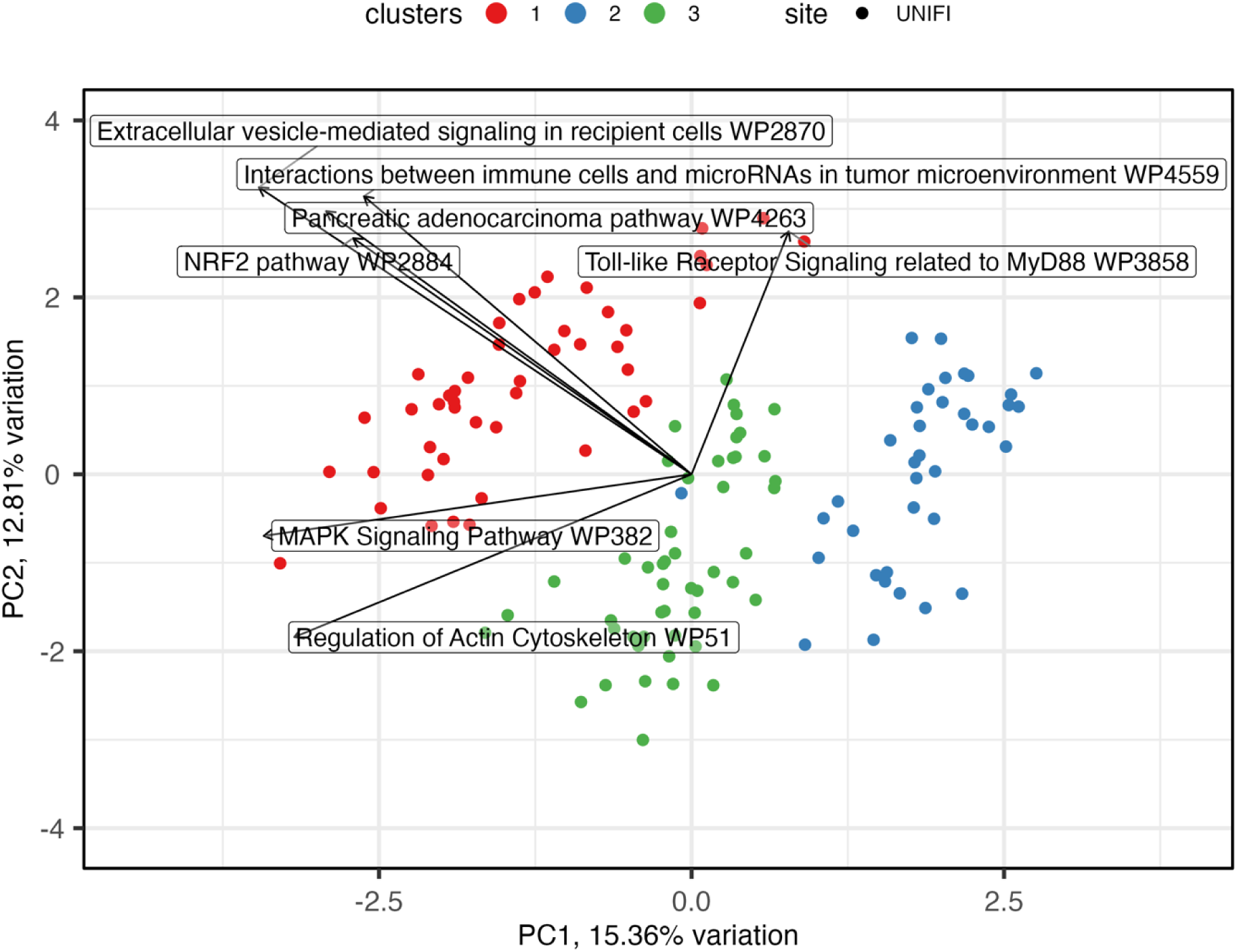
TopMD pathway volumes of each patient in the Florence cohort, plotted as a PCA plot. The data reveal three distinct clusters based on pathway activation, as determined by k-means.

### Molecular phenotypes and pathway activation

Cluster 1 was characterised by high activation of pathways related to ESC pluripotency, NRF2, and TGF-β receptor signalling (Figure 2). Cluster 2 showed elevated activity in focal adhesion– PI3K–Akt–mTOR and type I interferon signalling pathways. Cluster 3 displayed low activation of IRF7-related pathways.

**Figure 2:**
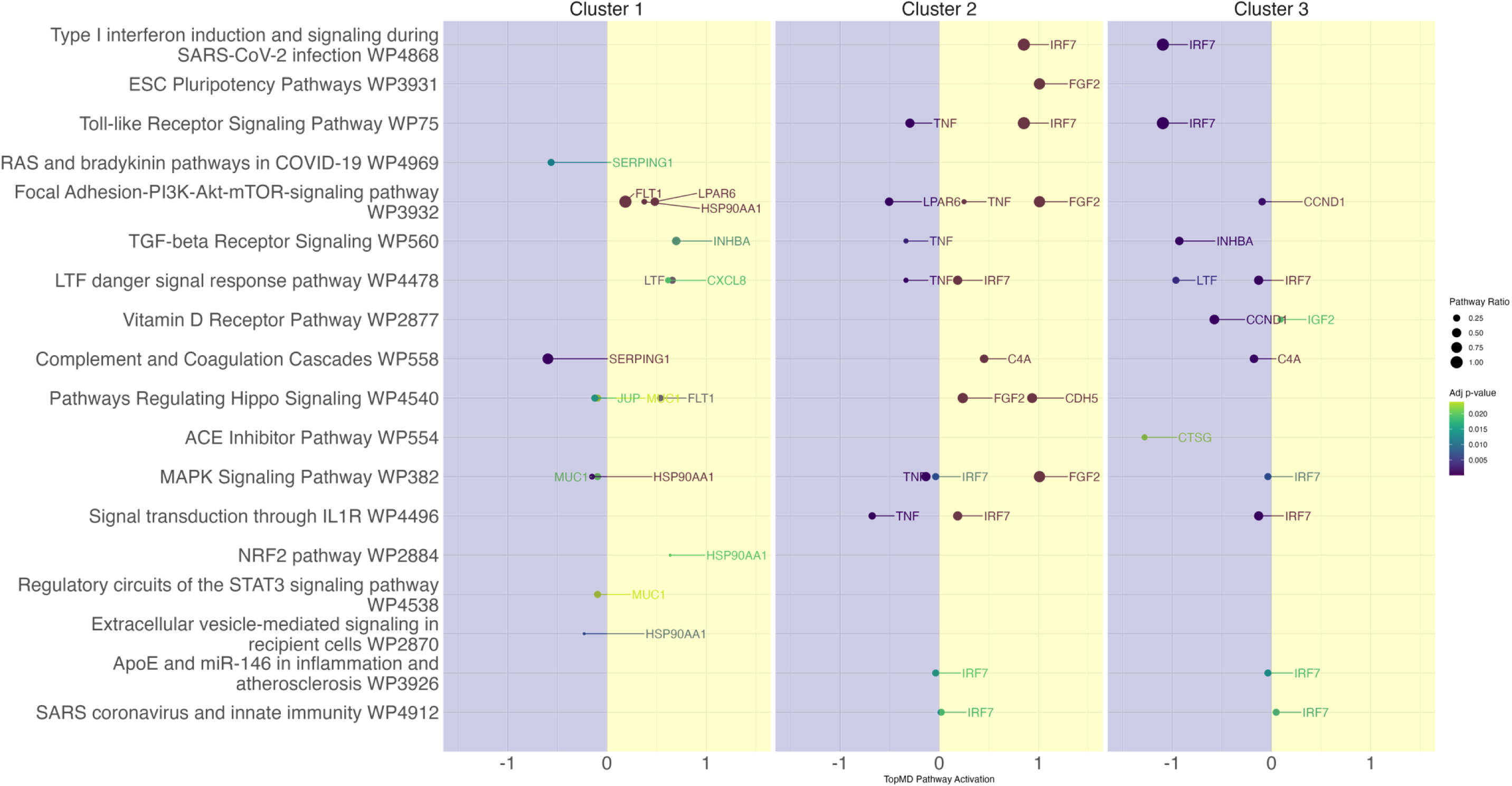
The average pathway volume for each cluster was considered in a TopMD enrichment analysis against the average pathway activation for the whole cohort to identify differentially activated pathways. The enrichment analysis was filtered by adjusted P value, then the top pathways were plotted. The pathways are annotated with the gene that leads the identified pathway. The dots are coloured by adjusted p-value, and the size represents the proportion of genes identified within that pathway from TopMD analysis.

### Classification performance and validation

LRROC analysis was conducted on models trained with 70% of Florence patients, with testing on the remaining 30%. AUCROC scores were 0.84, 0.85, and 0.72 for Clusters 1, 2, and 3, respectively. Validation in the Liège cohort yielded AUCROC values of 0.76, 0.93, and 0.69 for Clusters 1, 2, and 3, respectively (Supplementary Figures 1 and 2).

### Potential drug candidates for each cluster

TopMD pathway activation profiles were mapped to the Drug-Gene Interaction Database (Figure 3), identifying potential drug targets specific to each cluster (Supplementary Table 4). This dual-purpose approach provides insights into both potential therapeutic strategies and the underlying biology of each phenotype. While shared targets such as *ITGB2*, *GNAS*, and *CXCR2* appeared across all clusters, unique targets emerged: *IFNAR1*, *TGFBR2*, and *CSF2RB* in Cluster 1; *SERPING1* and *TLN1* in Cluster 2; and *SERPING1* (shared with Cluster 2) in Cluster 3. These distinctions emphasise both common and cluster-specific therapeutic opportunities.

**Figure 3:**
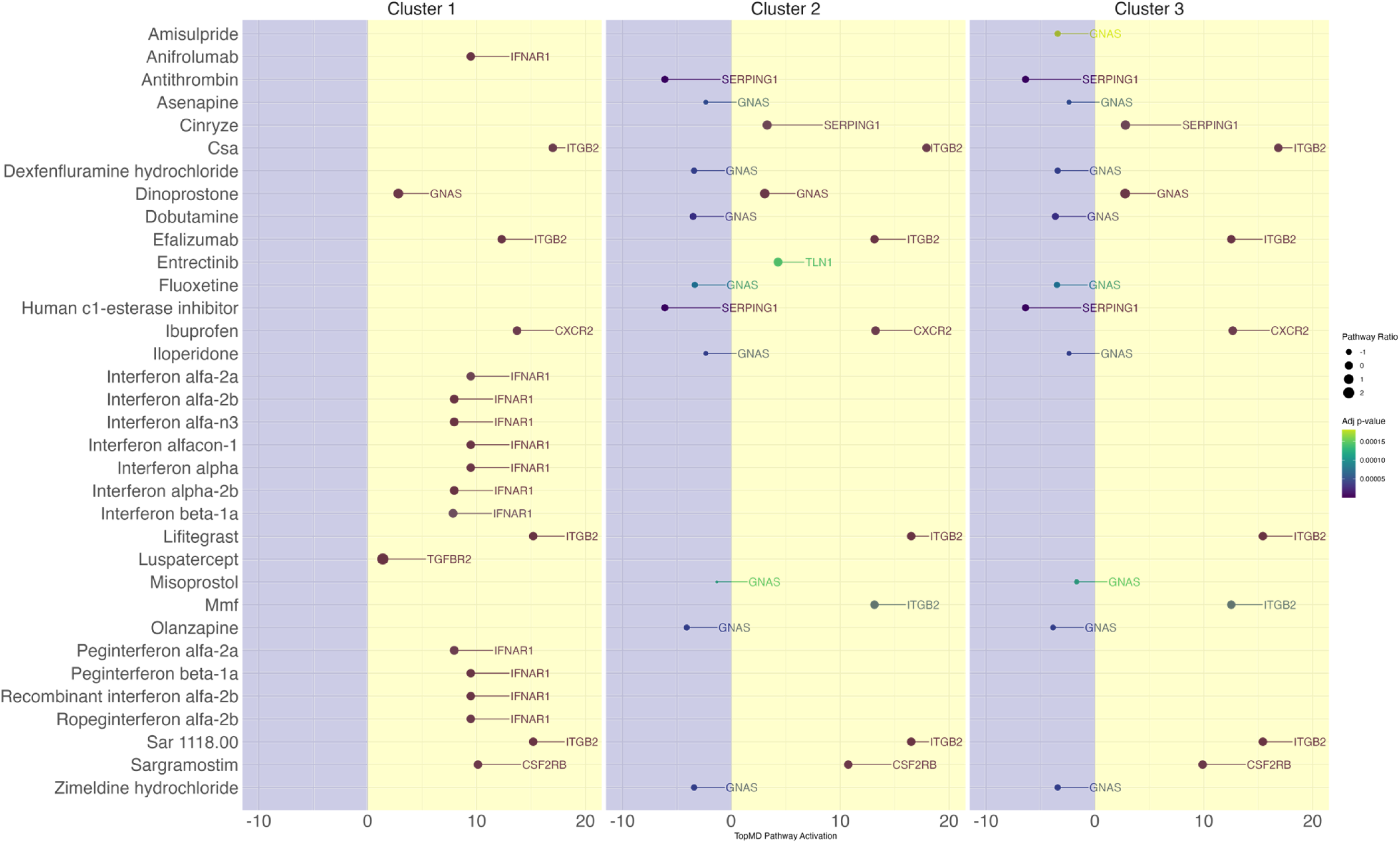
TopMD enrichment analysis was mapped against the Drug-Gene Interaction Database, using a healthy baseline, revealing approved drugs that are known to target genes and their corresponding pathways. The top drug candidates are plotted based on adjusted p-value and pathway volume.

### Pathway activation in fatal cases: identifying potential intervention targets

Pathway activation was investigated in the 10 deceased patients from the Florence and Liège cohorts. As expected, these patients were elderly and had high rates of comorbidities— cardiovascular (70%), respiratory (50%), malnutrition (40%), hypertension (90%), cerebrovascular (30%), and chronic hepatitis (40%). Notably, all 10 exhibited strong activation of the non-alcoholic fatty liver disease pathway, primarily driven by NDUFA9 and UQCRC2 (Figure 4).

**Figure 4:**
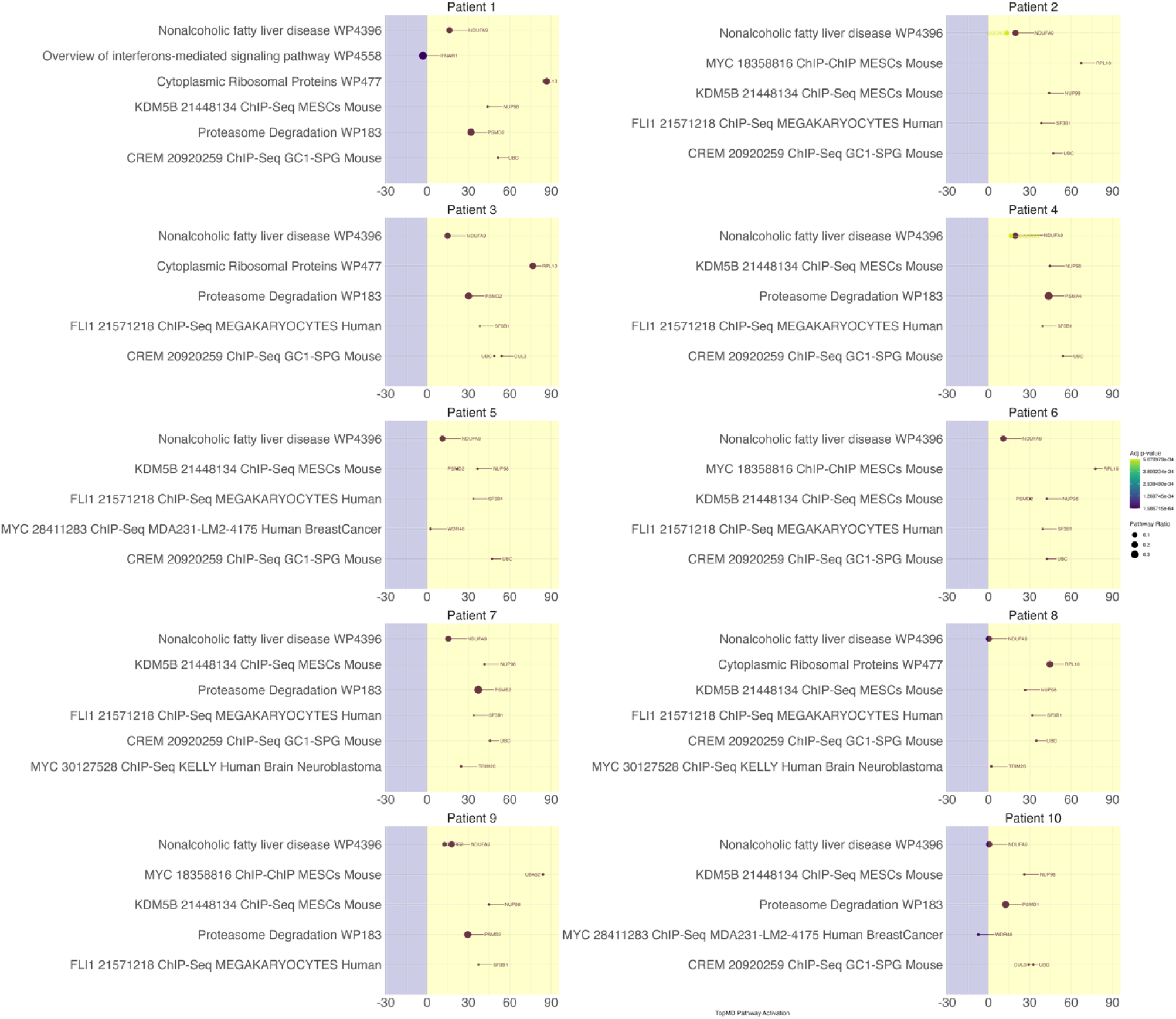
The top 6 pathways enriched in fatal cases within the Florence and Liège cohort using a healthy baseline.

Despite this shared pathway, analysis at the individual level revealed considerable heterogeneity, reflecting the complex interaction between COVID-19, comorbidities, and personal demographics. Enrichment analysis identified potential therapeutic targets based on individual pathway profiles. All patients showed activation of druggable targets such as CXCR2 (Figure 5), while additional targets were found in specific individuals, including GNAS (multiple patients), ITGB2 (patients 2 and 6), CSF2RB (multiple patients), SERPING1 (5 patients), PIK3CD (patient 5), TGFBR2 (patient 9), and CUL4B (patient 10).

**Figure 5:**
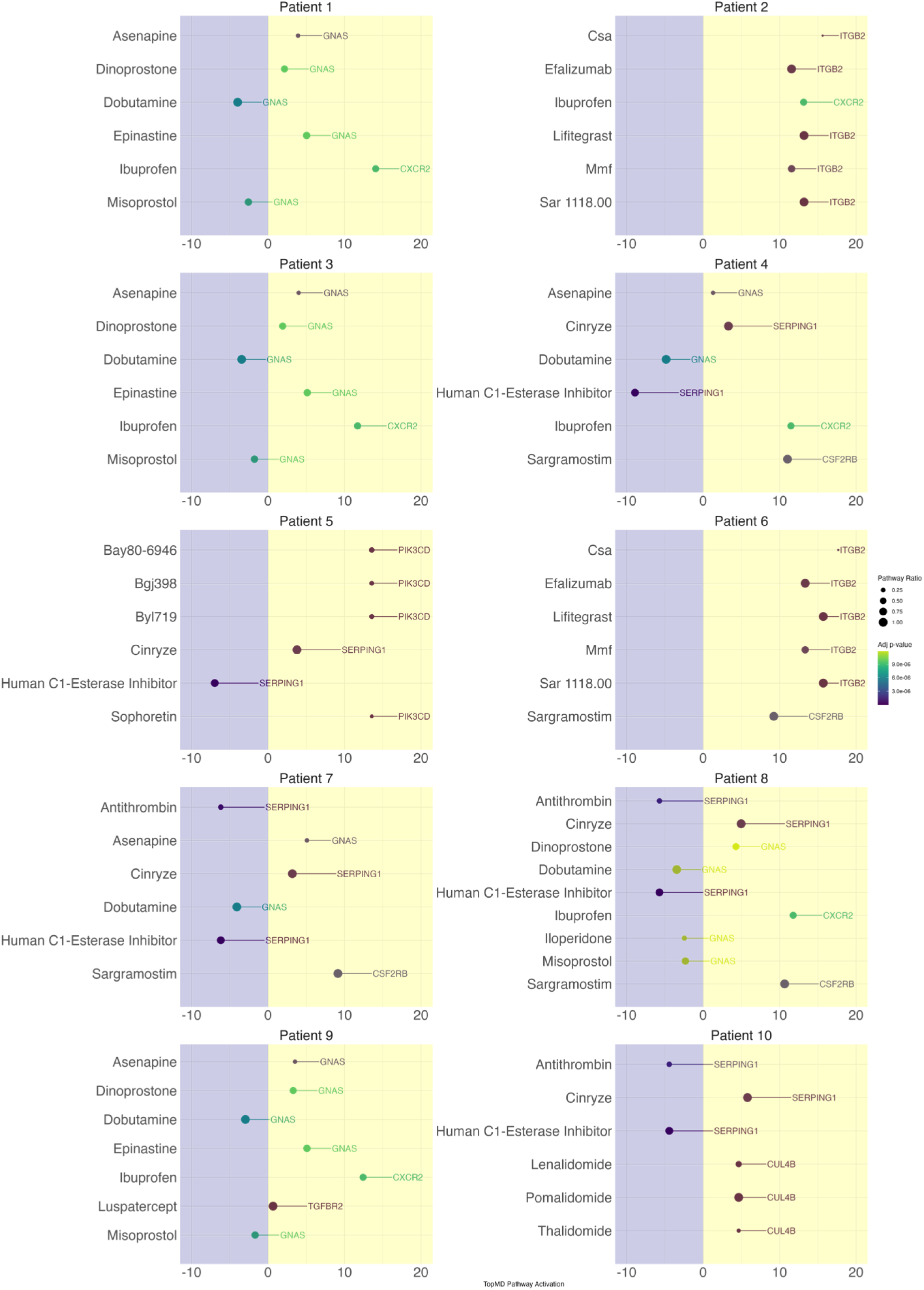
The top significant drug candidates and peak genes that could potentially modulate the phenotype of the 10 fatal cases patients in the Florence and Liège cohort.

## Discussion

Traditional molecular phenotyping relies on data reduction and feature selection to remove biological and technical ‘noise’ before pathway enrichment analysis. However, this can obscure essential signals. TopMD topological analysis overcomes this limitation by capturing global gene expression patterns, including low-abundance transcripts often discarded. These transcripts form the ‘foothills’ of highly activated pathways, contributing to a comprehensive view of the molecular phenotype. Understanding these phenotypes enables more precise therapeutic selection, as drugs act at the molecular rather than clinical level^22^.

There is increasing recognition of the need to integrate diverse data types—such as biomarkers, clinical measures, and imaging—to refine outcome prediction. AI frameworks applied to chest CT, for example, have achieved high diagnostic accuracy (85.2%) for COVID-19 and facilitated rapid clinical decision-making^23^. Another AI tool, CACOVID-CT, quantified lung damage severity and correlated with hospital stay, ICU admission, mechanical ventilation, and mortality across 476 patients^24^. These tools enhance risk stratification and reduce radiologist workload.

In our study, three molecular phenotypes of COVID-19 were identified via TopMD analysis of blood transcriptomics. These clusters showed strong classification performance in the Florence cohort (AUC: 0.84–0.85) and validated well in the Liège cohort (AUC: 0.69–0.93). Cluster-specific clinical features emerged: clusters 1 and 2 had elevated lactic acid, a known marker of severity^25^; cluster 2 also showed more respiratory disease, higher oxygen demand, and raised direct bilirubin^26^. Most COVID-19 fatalities occurred in cluster 2. Importantly, CORADS CT scores could not distinguish clusters, suggesting underlying molecular differences despite similar radiological findings. Continuous CT scoring may offer improved granularity^23^.

Cluster 1 was marked by high activation of ESC pluripotency, NRF2, and TGF-β receptor signalling—pathways linked to tissue repair, antioxidant response, and inflammation regulation^22,27^. Conversely, extracellular vesicle-mediated signalling and complement/coagulation cascades were underactive, suggesting impaired intercellular communication and immune dysregulation^28^. Cluster 2 showed increased PI3K-Akt-mTOR and type I interferon pathway activation, associated with antiviral responses and tissue repair^29,30^. Cluster 3 exhibited low IRF7, TGF-β, and IL-1R signalling, as well as a reduced LTF danger signal response. Diminished lactoferrin signalling may impair viral control and modulate inflammation^31,32^.

Drug-pathway mapping using the Drug-Gene Interaction Database revealed both shared and unique therapeutic targets across clusters (Figure 3, Supplementary Table 4). For example, ITGB2, GNAS, and CXCR2 were common targets, while IFNAR1, TGFBR2, and CSF2RB were specific to Cluster 1. Cluster 2 included SERPING1, TLN1, and Cluster 3 shared SERPING1. This offers potential for cluster-specific repurposing of existing therapies.

Cyclosporine A (CSA) emerged as a candidate across all clusters, having shown safety and potential to reduce hyperinflammation in COVID-19^33,34^. Interferon-modulating drugs and TGF-β inhibitors such as Luspatercept were identified in Cluster 1, aligning with reports of their efficacy^22,35^. Lifitegrast, shown to inhibit SARS-CoV-2 in vitro^36^, also matched this cluster. In Cluster 2, SSRIs like fluoxetine and fluvoxamine were highlighted, in line with evidence suggesting benefit in COVID-19 and long-COVID^37^. Cinryze (C1 esterase inhibitor) and Asenapine were identified as cluster-specific candidates^38,39^. Amisulpride emerged as a target in Cluster^3^.

### Individual-level precision medicine in fatal cases

Enrichment analysis of the 10 deceased patients (Florence and Liège) further demonstrated TopMD’s utility in individualised treatment guidance. While all patients shared CXCR2 and GNAS activation (suggesting potential benefit from drugs like ibuprofen), other targets varied. For example, ITGB2 was enriched in patients 2 and 6, CSF2RB in several patients (implicating Sargramostim)^40^, and SERPING1 in five patients (suggesting use of Cinryze or antithrombin). Patient-specific targets included PIK3CD (patient 5; candidate: Sophoretin)^28^, TGFBR2 (patient 9; candidate: Luspatercept), and CUL4B (patient 10; candidate: Thalidomide, Pomalidomide, or Lenalidomide)^41,42^.

This research identified three molecular phenotypes in hospitalised COVID-19 patients, which were not reflected in CT imaging or routine clinical observations. These phenotypes correspond to distinct molecular mechanisms and drug targets, supporting a biomarker-based, stratified approach to treatment.

Topological analysis of global gene expression to define a patient’s pathway activation map could be used in future pandemics to assist treatment decisions before clinical trials are completed and to identify and prioritise candidate therapeutics for rapid testing.

**Table 1:**
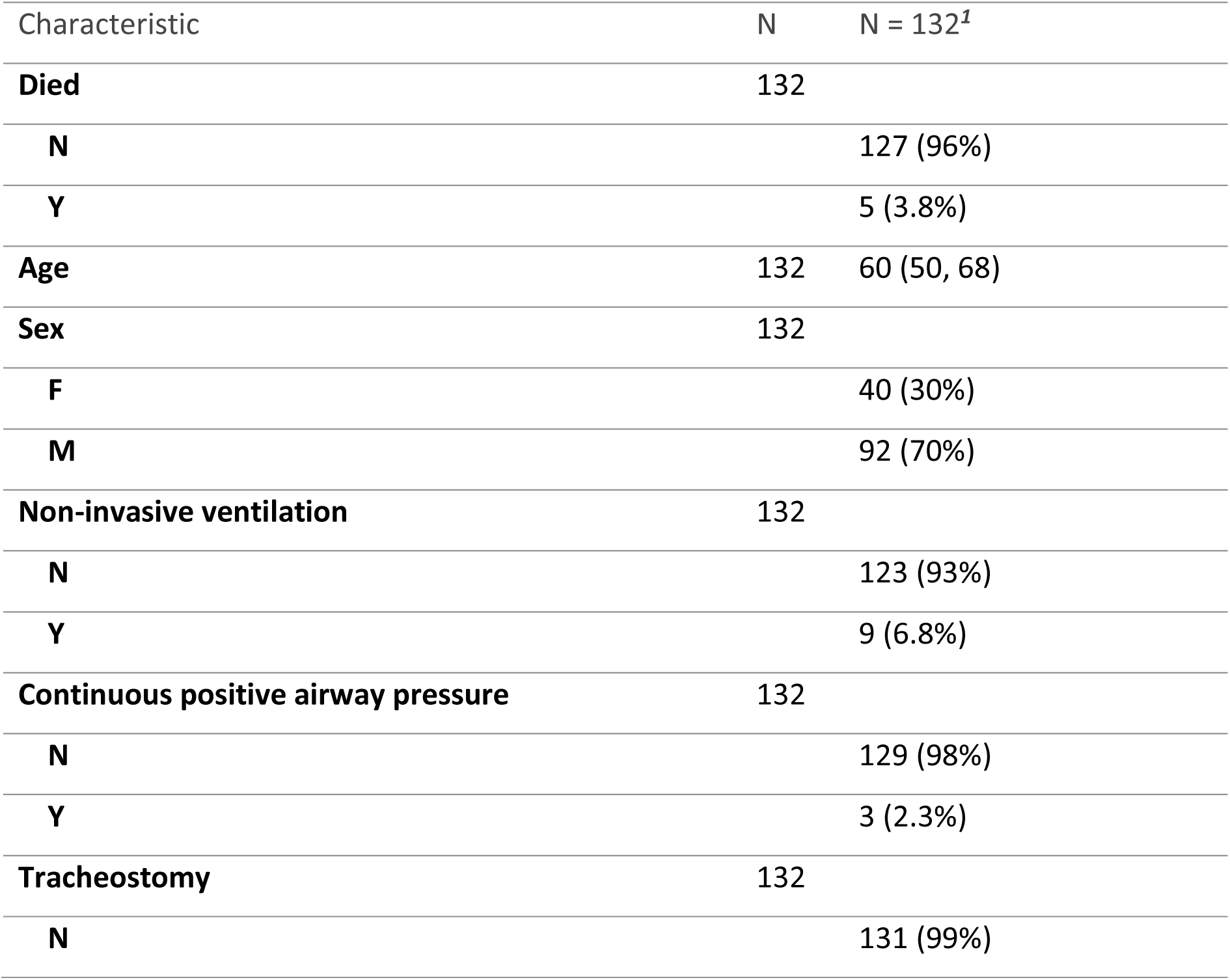
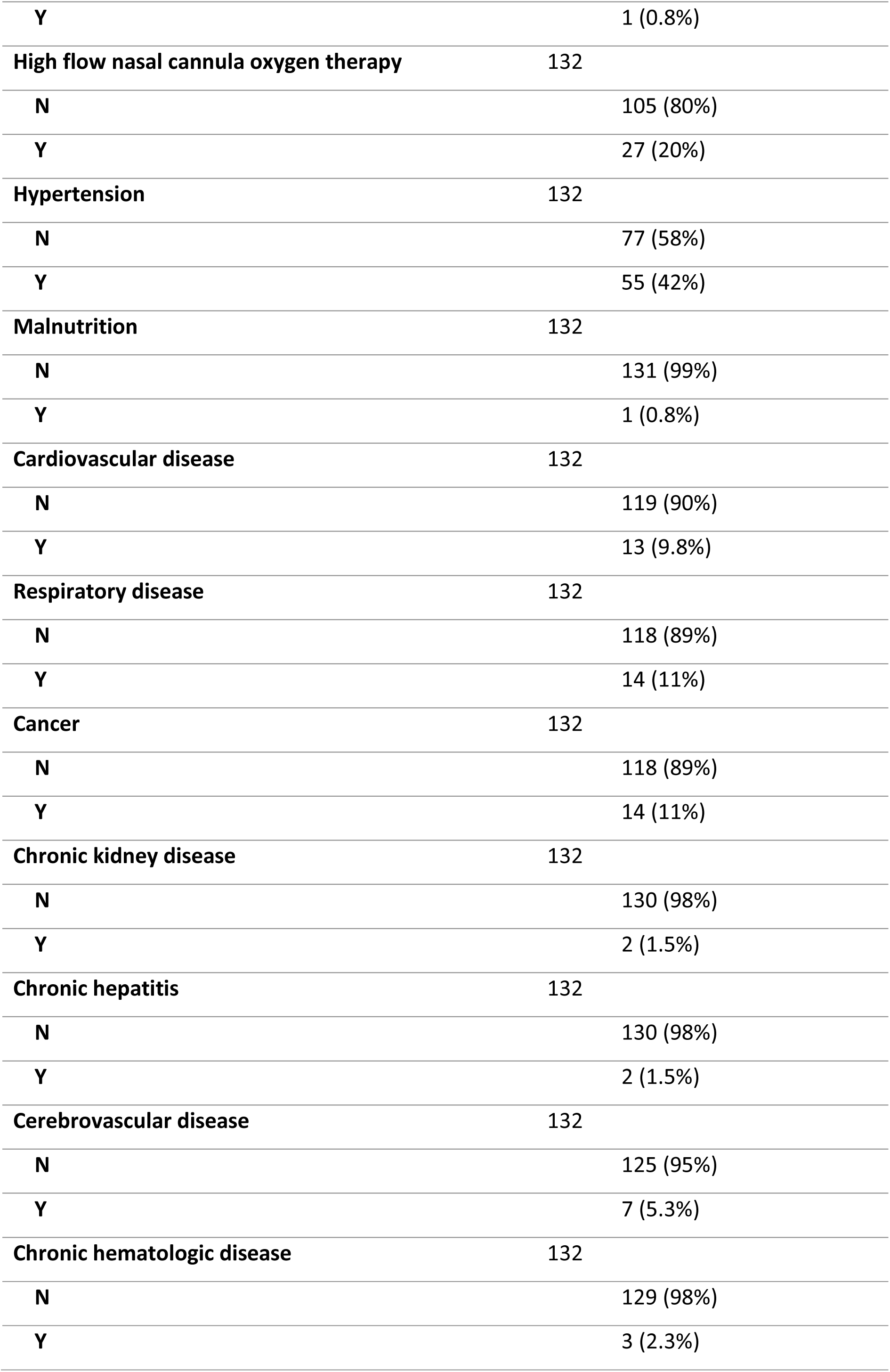
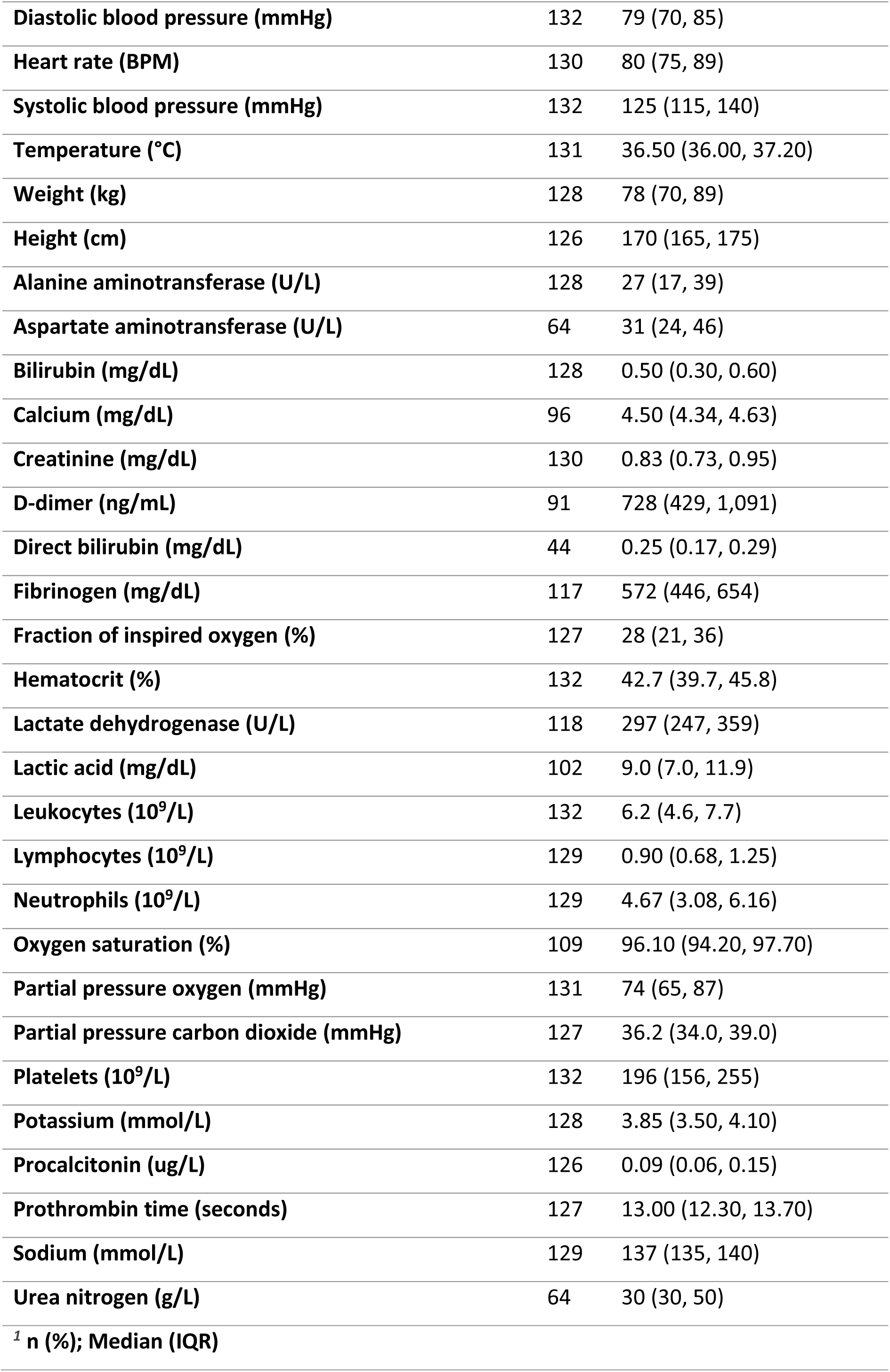
Characteristics of 132 patients from Florence included in the study, including lab results at admission.

**Table 2:** Patient characteristics that differ between the three clusters in the Florence cohort (p = <0.05).

| Characteristic | N | 1, N = 46<sup>1</sup> | 2, N = 37<sup>1</sup> | 3, N = 49<sup>1</sup> | p-value<sup>2</sup> |
| --- | --- | --- | --- | --- | --- |
| Lactic acid (mg/dL) | 102 | 10.0 (7.7, 13.0) | 10.0 (7.2, 12.0) | 8.0 (5.3, 9.7) | 0.008 |
| Fraction of inspired oxygen (%) | 127 | 28 (21, 36) | 32 (27, 40) | 28 (21, 29) | 0.019 |
| Died | 132 |  |  |  | 0.032 |
| N |  | 46 (100%) | 33 (89%) | 48 (98%) |  |
| Y |  | 0 (0%) | 4 (11%) | 1 (2.0%) |  |
| Respiratory disease | 132 |  |  |  | 0.042 |
| N |  | 44 (96%) | 29 (78%) | 45 (92%) |  |
| Y |  | 2 (4.3%) | 8 (22%) | 4 (8.2%) |  |
| Direct bilirubin (mg/dL) | 44 | 0.20 (0.17, 0.27) | 0.28 (0.24, 0.32) | 0.20 (0.17, 0.28) | 0.047 |

<sup>1</sup> n (%); Median (IQR)
<sup>2</sup> Fisher's exact test; Kruskal-Wallis rank sum test; Pearson's Chi-squared test

## Acknowledgments

The authors wish to thank the study participants and the hospital staff for their participation in this study.

## Contributions

RPR, FS: methodology, RPR, FS: visualisation, RPR, FS, BVE, TK: software, BE: project administration, RPR, FS, JG, MH, AS, EG, LG, AJP, CN: investigation, RPR, FS, JG, CH, TP, SW: formal analysis, JG, CN: clinical supervision, RPR, FS, AJP, ST, RB, KH, XX, YN, SW: data curation, RPR, FS, EP, PS, JAH, JPS: writing - original draft preparation, RPR, FB, BE, BE, JG, MH, AS, EG, LG, AJP, CN, ST, RB, KH, CH, TP, RD, TC, DB, SW, XX, YN, SW, SW, GY, PJS, JAH, JPRS: writing - review and editing, JPRS, JAH, PJS, GY, SW: funding acquisition

## Data availability

Sequencing reads available under SRA bioproject: PRJNA1085259

