## Supplementary Material for "Precision medicine for pandemics: stratification of COVID-19 molecular phenotypes defined by topological analysis of global blood gene expression"

**Supplementary figures**

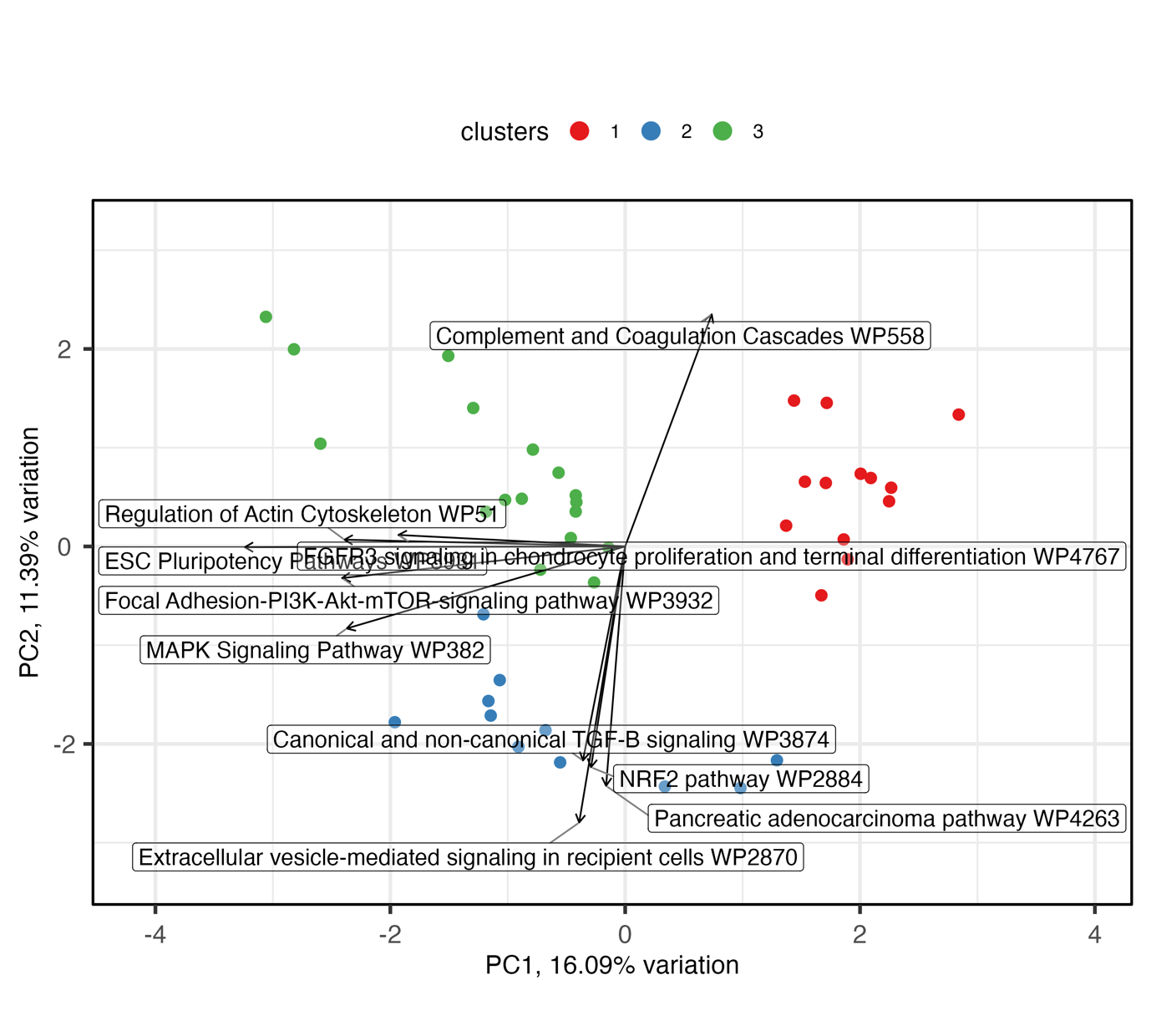

**Supplementary Figure 1:** PCA plot of pathway volumes from the Liège validation cohort.

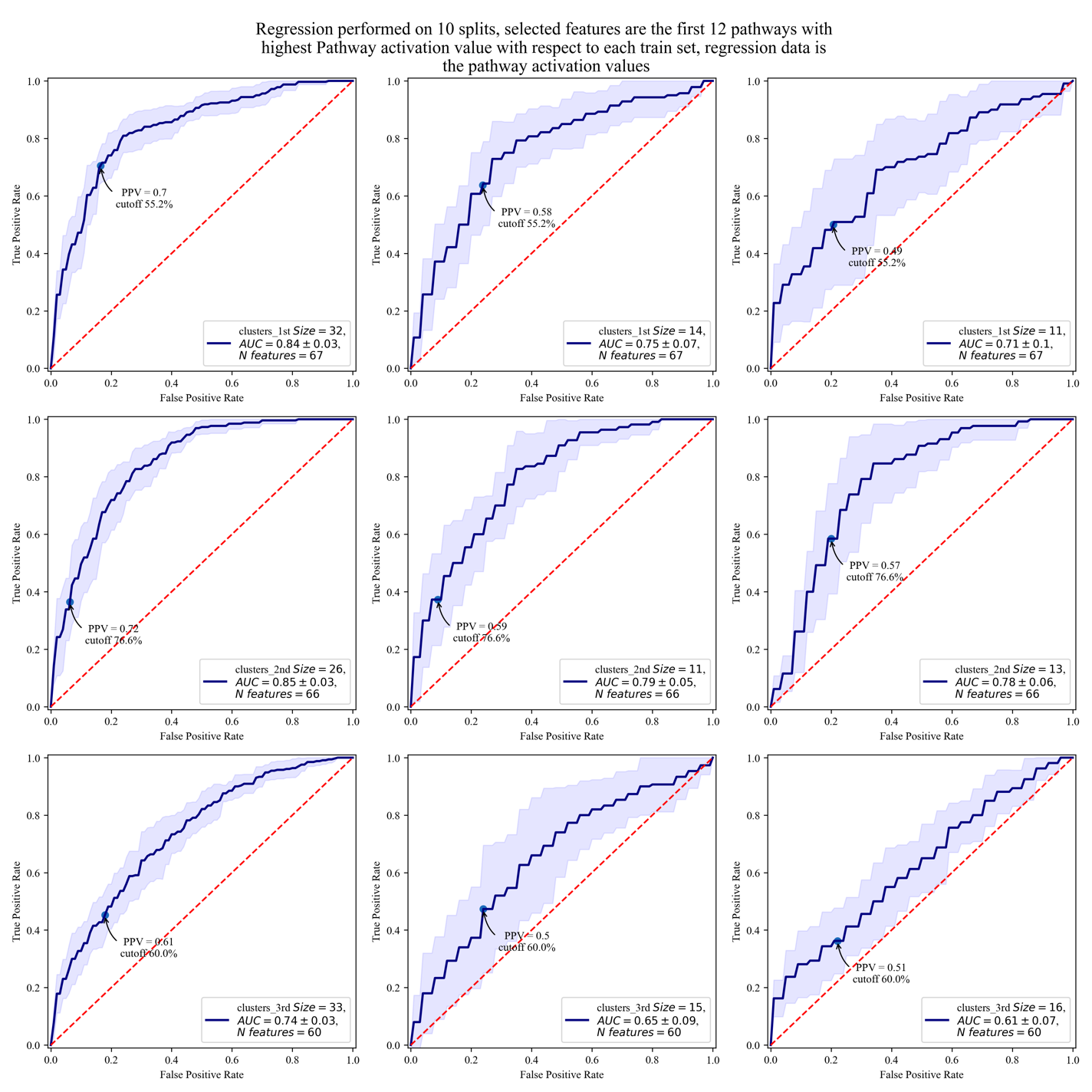

**Supplementary Figure 2:**

ROC curves for the different clusters, using the same parameters, in this case, selecting the **N** highest pathways in term of activation volume (squared sum of Log2 Fold Change of the pathways) and computing the regression model on the matrix, which entries were such activation volumes per each sample, created by TopMD. First and second columns contain ROC curves for the 10 pair of training and test sets, balanced in terms of the classes, such that the union of each pair is the whole Unifi dataset. The third column instead represents the ROC curves of the predictions of such models on the LIEGE dataset. An optimal cutoff value for classification from LR prediction was computed for each class, using the slope of the ROC curves on different cutoff values.

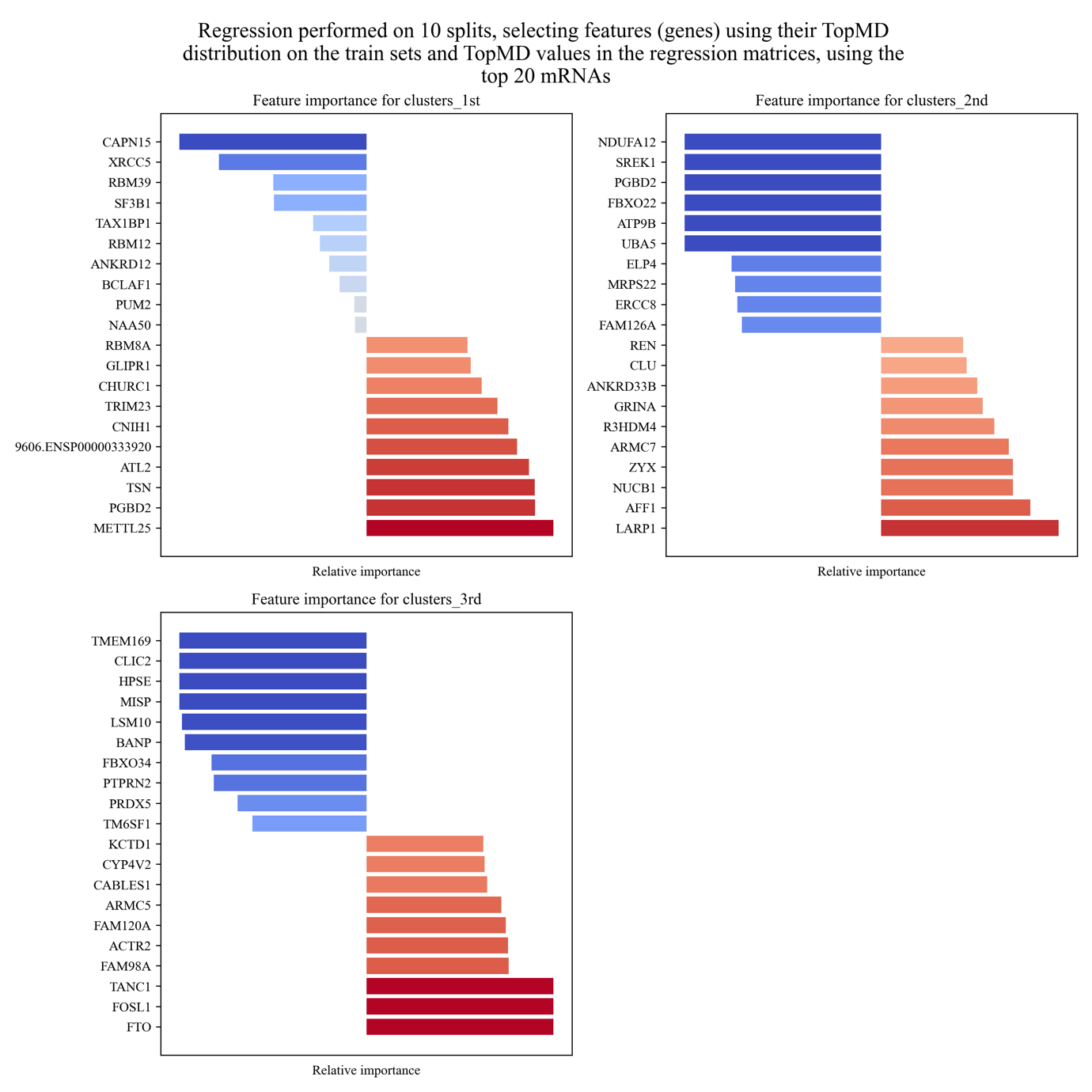

**Supplementary Figure 3:**

Average relative importance of each variable used in the regression models in Figure 2, only the 10 highest and 10 lowest, when present are plotted. Colour ranges from red (high and positive) to blue (low and negative) depending on the average relative importance.

**Supplementary Tables**

**Supplementary Table 1: CT scan observations from Florence and Liège cohorts**

| Characteristic | N | N = 109*^1^* |
| --- | --- | --- |
| COVID-19 Diagnosis based on CORADS | 109 |  |
| 1: Very Low |  | 3 (2.8%) |
| 2: Low |  | 5 (4.6%) |
| 3: Equivocal/unsure |  | 28 (26%) |
| 4: High |  | 44 (40%) |
| 5: Very High |  | 29 (27%) |
| LUL | 109 |  |
| 0: 0% |  | 3 (2.8%) |
| 1: 0-5% |  | 18 (17%) |
| 2: 5-25% |  | 46 (42%) |
| 3: 25-50% |  | 24 (22%) |
| 4: 50-75% |  | 15 (14%) |
| 5: 75%+ |  | 3 (2.8%) |
| LLL | 109 |  |
| 0: 0% |  | 4 (3.7%) |
| 1: 0-5% |  | 5 (4.6%) |
| 2: 5-25% |  | 39 (36%) |
| 3: 25-50% |  | 28 (26%) |
| 4: 50-75% |  | 17 (16%) |
| 5: 75%+ |  | 16 (15%) |
| RUL | 109 |  |
| 0: 0% |  | 8 (7.3%) |
| 1: 0-5% |  | 17 (16%) |
| 2: 5-25% |  | 46 (42%) |
| 3: 25-50% |  | 16 (15%) |
| 4: 50-75% |  | 15 (14%) |
| 5: 75%+ |  | 7 (6.4%) |
| RML | 109 |  |
| 0: 0% |  | 7 (6.4%) |
| 1: 0-5% |  | 21 (19%) |
| 2: 5-25% |  | 47 (43%) |
| 3: 25-50% |  | 19 (17%) |
| 4: 50-75% |  | 9 (8.3%) |
| 5: 75%+ |  | 6 (5.5%) |
| RLL | 109 |  |
| 0: 0% |  | 2 (1.8%) |
| 1: 0-5% |  | 8 (7.3%) |
| 2: 5-25% |  | 31 (28%) |
| 3: 25-50% |  | 40 (37%) |
| 4: 50-75% |  | 16 (15%) |
| 5: 75%+ |  | 12 (11%) |
| Patterns | 133 |  |
| Crazy paving |  | 13 (9.8%) |
| Emphysema |  | 3 (2.3%) |
| GGO and consolidation |  | 80 (60%) |
| Multi-focal |  | 8 (6.0%) |
| None |  | 1 (0.8%) |
| Only consolidation |  | 3 (2.3%) |
| Only GGO |  | 25 (19%) |
| Distributions | 152 |  |
| Apical |  | 2 (1.3%) |
| Bronchocentric |  | 14 (9.2%) |
| Diffuse |  | 36 (24%) |
| Dorsal |  | 14 (9.2%) |
| Lower lobes |  | 29 (19%) |
| Mantle |  | 1 (0.7%) |
| None |  | 1 (0.7%) |
| Peripheral |  | 54 (36%) |
| Right lung |  | 1 (0.7%) |
| Additional COVID-19 findings | 187 |  |
| Halo sign |  | 5 (2.7%) |
| None |  | 36 (19%) |
| Perilobular pattern (OP signs) |  | 9 (4.8%) |
| Pleural thickening |  | 27 (14%) |
| Subpleural lines |  | 49 (26%) |
| Subpleural sparing |  | 19 (10%) |
| Vascular enlargement |  | 42 (22%) |
| Contra-indicative findings | 134 |  |
| Atelectasis |  | 10 (7.5%) |
| Discrete small nodules |  | 3 (2.2%) |
| Isolated lobar/segmental consolidation |  | 6 (4.5%) |
| Lymphadenopathy |  | 29 (22%) |
| None |  | 59 (44%) |
| Pericardial effusion |  | 3 (2.2%) |
| Pleural effusion |  | 12 (9.0%) |
| Smooth interlobular septal thickening |  | 9 (6.7%) |
| Tree in bud |  | 3 (2.2%) |
| Comorbidities | 125 |  |
| Ascites |  | 1 (0.8%) |
| Aortic/coronaric calcifications |  | 48 (38%) |
| Cardiomegaly |  | 7 (5.6%) |
| Hepatic calcification |  | 1 (0.8%) |
| None |  | 47 (38%) |
| Oncologic Pt |  | 2 (1.6%) |
| Pre-existing pulmonary disease |  | 5 (4.0%) |
| Pulmonary hypertension |  | 1 (0.8%) |
| Steatosis |  | 11 (8.8%) |
| Tracheomalacia |  | 2 (1.6%) |
| *^1^* n (%) | | |

**Supplementary Table 2:** Characteristics of 41 patients from Liège included in the study, including lab results at admission.

| **Characteristic** | **N** | **N = 41***^1^* |
| --- | --- | --- |
| Died | 41 |  |
| N |  | 36 (88%) |
| Y |  | 5 (12%) |
| Age | 41 | 70 (61, 75) |
| Sex | 41 |  |
| F |  | 15 (37%) |
| M |  | 26 (63%) |
| Non-invasive ventilation | 41 |  |
| N |  | 38 (93%) |
| Y |  | 3 (7.3%) |
| Continuous positive airway pressure | 41 |  |
| N |  | 41 (100%) |
| Tracheostomy | 41 |  |
| N |  | 41 (100%) |
| High flow nasal cannula oxygen therapy | 41 |  |
| N |  | 32 (78%) |
| Y |  | 9 (22%) |
| Mechanical ventilation | 3 |  |
| Y |  | 3 (100%) |
| Hypertension | 41 |  |
| N |  | 9 (22%) |
| Y |  | 32 (78%) |
| Malnutrition | 41 |  |
| N |  | 22 (54%) |
| Y |  | 19 (46%) |
| Cardiovascular disease | 41 |  |
| N |  | 9 (22%) |
| Y |  | 32 (78%) |
| Respiratory disease | 41 |  |
| N |  | 28 (68%) |
| Y |  | 13 (32%) |
| Cancer | 41 |  |
| N |  | 36 (88%) |
| Y |  | 5 (12%) |
| Chronic kidney disease | 41 |  |
| N |  | 32 (78%) |
| Y |  | 9 (22%) |
| Chronic hepatitis | 41 |  |
| N |  | 18 (44%) |
| Y |  | 23 (56%) |
| Cerebrovascular disease | 41 |  |
| N |  | 36 (88%) |
| Y |  | 5 (12%) |
| Chronic hematologic disease | 41 |  |
| N |  | 40 (98%) |
| Y |  | 1 (2.4%) |
| Diastolic blood pressure (mmHg) | 41 | 77 (64, 82) |
| Heart rate (BPM) | 41 | 96 (81, 99) |
| Systolic blood pressure (mmHg) | 41 | 130 (122, 143) |
| Temperature (°C) | 41 | 36.20 (36.00, 36.70) |
| Weight (kg) | 27 | 75 (64, 87) |
| Height (cm) | 23 | 170 (163, 176) |
| Alanine aminotransferase (U/L) | 41 | 30 (21, 44) |
| Albumin (g/L) | 41 | 38.0 (36.0, 40.0) |
| Aspartate aminotransferase (U/L) | 41 | 44 (31, 73) |
| Bilirubin (mg/dL) | 41 | 0.65 (0.52, 1.05) |
| Calcium (mmol/L) | 41 | 2.16 (2.07, 2.24) |
| Creatine kinase mb (U/L) | 41 | 105 (74, 146) |
| Creatinine (mg/dL) | 41 | 1.02 (0.78, 1.17) |
| D-dimer (ug/L) | 32 | 912 (499, 1,728) |
| Direct bilirubin (mg/dL) | 41 | 0.29 (0.20, 0.43) |
| Fibrinogen (g/L) | 41 | 574 (457, 702) |
| Hematocrit (%) | 41 | 39.9 (36.4, 42.8) |
| Lactate dehydrogenase (U/L) | 41 | 370 (278, 475) |
| Leukocytes (10^3^/mm^3^) | 41 | 6.4 (4.6, 8.8) |
| Lymphocytes (10^3^/mm^3^) | 41 | 0.83 (0.68, 1.01) |
| Neutrophils (10^3^/mm^3^) | 41 | 4.74 (3.09, 7.22) |
| Oxygen saturation (%) | 41 | 92.0 (91.0, 96.0) |
| Partial pressure oxygen (mmHg) | 25 | 66 (57, 74) |
| Partial pressure carbon dioxide (mmHg) | 25 | 31 (30, 37) |
| Platelets (10^3^/mm^3^) | 41 | 171 (150, 242) |
| Potassium (mmol/L) | 41 | 4.02 (3.64, 4.48) |
| Procalcitonin (ug/L) | 36 | 0.14 (0.06, 0.38) |
| Prothrombin intl normalized ratio | 41 | 1.06 (1.01, 1.15) |
| Prothrombin time (seconds) | 41 | 12.40 (11.90, 13.50) |
| Sodium (mmol/L) | 41 | 138 (136, 141) |
| Urea nitrogen (mg/dL) | 41 | 45 (32, 54) |
| *^1^* n (%); Median (IQR) | | |

**Supplementary Table 3: Significant differences observed between the 3 molecular phenotypes in the Liège cohort.**

| **Characteristic** | **N** | **1**, N = 11*^1^* | **2**, N = 17*^1^* | **3**, N = 13*^1^* | **p-value***^2^* |
| --- | --- | --- | --- | --- | --- |
| **Potassium (mmol/L)** | 41 | 3.64 (3.35, 3.90) | 4.34 (4.13, 4.59) | 3.80 (3.70, 4.33) | 0.017 |
| *^1^* n (%); Median (IQR) | | | | | |
| *^2^* Fisher’s exact test; Kruskal-Wallis rank sum test | | | | | |

**Supplementary table 4:** FDA approved drugs identified as modulators of pathway activity in each cluster identified in the UNIFI cohort.

| Drug Name | Cluster 1 | Cluster 2 | Cluster 3 |
| --- | --- | --- | --- |
| 13-cis-retinoic acid | 🗸 |  |  |
| 17 beta-estradiol | 🗸 |  |  |
| 2-cda | 🗸 |  |  |
| 4-hpr | 🗸 |  |  |
| 5-fluorouracil | 🗸 | 🗸 | 🗸 |
| 5-fu | 🗸 | 🗸 | 🗸 |
| Abciximab |  | 🗸 |  |
| Acalabrutinib | 🗸 | 🗸 | 🗸 |
| Acalabrutinib maleate | 🗸 | 🗸 | 🗸 |
| Acebutolol | 🗸 | 🗸 | 🗸 |
| Acebutolol hydrochloride | 🗸 | 🗸 | 🗸 |
| Acetaminophen | 🗸 | 🗸 | 🗸 |
| Acetophenazine | 🗸 | 🗸 | 🗸 |
| Acp-196 | 🗸 | 🗸 | 🗸 |
| Adalimumab | 🗸 | 🗸 | 🗸 |
| Adenosine | 🗸 | 🗸 | 🗸 |
| Adenosine monophosphate | 🗸 | 🗸 | 🗸 |
| Adriamycin | 🗸 |  |  |
| Ajmalicine | 🗸 | 🗸 | 🗸 |
| Albuterol |  | 🗸 |  |
| Albuterol sulfate |  | 🗸 |  |
| Alpelisib | 🗸 |  | 🗸 |
| Alpha-methyl-p-tyrosine | 🗸 |  |  |
| Alprazolam | 🗸 | 🗸 | 🗸 |
| Alprostadil | 🗸 | 🗸 | 🗸 |
| Alum | 🗸 | 🗸 | 🗸 |
| Aluminum hydroxide | 🗸 | 🗸 | 🗸 |
| Amg 416 | 🗸 | 🗸 | 🗸 |
| Amifostine | 🗸 |  |  |
| Amikacin | 🗸 | 🗸 | 🗸 |
| Amineptine | 🗸 | 🗸 | 🗸 |
| Aminophylline | 🗸 | 🗸 | 🗸 |
| Amiodarone | 🗸 | 🗸 | 🗸 |
| Amisulpride | 🗸 | 🗸 | 🗸 |
| Amitriptyline | 🗸 | 🗸 | 🗸 |
| Amitriptyline hydrochloride | 🗸 | 🗸 | 🗸 |
| Amoxapine | 🗸 | 🗸 | 🗸 |
| Amsacrine | 🗸 | 🗸 | 🗸 |
| Anakinra | 🗸 | 🗸 | 🗸 |
| Anastrozole |  |  | 🗸 |
| Anifrolumab | 🗸 |  |  |
| Antihistamine | 🗸 | 🗸 | 🗸 |
| Antithrombin |  | 🗸 | 🗸 |
| Antithymocyte globulin | 🗸 | 🗸 | 🗸 |
| Apomorphine | 🗸 | 🗸 | 🗸 |
| Aprotinin |  | 🗸 | 🗸 |
| Ara-c | 🗸 | 🗸 | 🗸 |
| Arbutamine | 🗸 | 🗸 | 🗸 |
| Arformoterol |  | 🗸 |  |
| Arformoterol tartrate |  | 🗸 |  |
| Asenapine | 🗸 | 🗸 | 🗸 |
| Aspirin | 🗸 | 🗸 | 🗸 |
| Atenolol | 🗸 | 🗸 | 🗸 |
| Atezolizumab | 🗸 |  | 🗸 |
| Atomoxetine | 🗸 | 🗸 | 🗸 |
| Atorvastatin | 🗸 |  | 🗸 |
| Atra | 🗸 | 🗸 | 🗸 |
| Beclomethasone dipropionate | 🗸 | 🗸 | 🗸 |
| Benzbromarone | 🗸 | 🗸 | 🗸 |
| Berotralstat hydrochloride |  | 🗸 | 🗸 |
| Betamethasone | 🗸 | 🗸 | 🗸 |
| Betaxolol | 🗸 | 🗸 | 🗸 |
| Betaxolol hydrochloride | 🗸 | 🗸 | 🗸 |
| Bethanidine | 🗸 | 🗸 | 🗸 |
| Bevacizumab | 🗸 | 🗸 | 🗸 |
| Bgb-3111 | 🗸 | 🗸 | 🗸 |
| Bgj398 | 🗸 |  | 🗸 |
| Bicalutamide | 🗸 | 🗸 | 🗸 |
| Bimatoprost | 🗸 | 🗸 | 🗸 |
| Bimekizumab | 🗸 | 🗸 | 🗸 |
| Bismuth subgallate |  | 🗸 | 🗸 |
| Bisoprolol | 🗸 | 🗸 | 🗸 |
| Bisoprolol fumarate | 🗸 | 🗸 | 🗸 |
| Bitolterol mesylate |  | 🗸 |  |
| Bms-986165 | 🗸 |  |  |
| Bortezomib | 🗸 | 🗸 | 🗸 |
| Bosutinib |  | 🗸 |  |
| Bretylium | 🗸 | 🗸 | 🗸 |
| Brodalumab | 🗸 | 🗸 | 🗸 |
| Bromocriptine | 🗸 | 🗸 | 🗸 |
| Bunolol | 🗸 | 🗸 | 🗸 |
| Buprenorphine |  | 🗸 |  |
| Bupropion | 🗸 | 🗸 | 🗸 |
| Busulfan | 🗸 | 🗸 | 🗸 |
| C1-inh |  | 🗸 | 🗸 |
| Cabozantinib | 🗸 | 🗸 | 🗸 |
| Caffeine | 🗸 |  | 🗸 |
| Caffeine, citrated | 🗸 | 🗸 | 🗸 |
| Calcitriol | 🗸 |  |  |
| Camptothecin | 🗸 | 🗸 | 🗸 |
| Carboplatin | 🗸 | 🗸 | 🗸 |
| Carphenazine | 🗸 | 🗸 | 🗸 |
| Carteolol | 🗸 | 🗸 | 🗸 |
| Carteolol hydrochloride | 🗸 | 🗸 | 🗸 |
| Carvedilol | 🗸 | 🗸 | 🗸 |
| Carvedilol phosphate | 🗸 | 🗸 | 🗸 |
| Ceftriaxone | 🗸 | 🗸 | 🗸 |
| Celiprolol | 🗸 | 🗸 | 🗸 |
| Cetirizine | 🗸 | 🗸 | 🗸 |
| Cetuximab | 🗸 | 🗸 | 🗸 |
| Chlorpromazine | 🗸 | 🗸 | 🗸 |
| Chlorprothixene | 🗸 | 🗸 | 🗸 |
| Cidofovir | 🗸 | 🗸 | 🗸 |
| Cilostazol | 🗸 | 🗸 | 🗸 |
| Cinacalcet | 🗸 | 🗸 | 🗸 |
| Cinacalcet hydrochloride | 🗸 | 🗸 | 🗸 |
| Cinryze |  | 🗸 | 🗸 |
| Ciprofloxacin | 🗸 | 🗸 | 🗸 |
| Cisapride | 🗸 | 🗸 | 🗸 |
| Cisplatin | 🗸 | 🗸 | 🗸 |
| Clarithromycin | 🗸 | 🗸 | 🗸 |
| Clebopride | 🗸 | 🗸 | 🗸 |
| Clenbuterol |  | 🗸 |  |
| Clenbuterol hydrochloride |  | 🗸 |  |
| Clobetasol propionate | 🗸 | 🗸 | 🗸 |
| Clomipramine | 🗸 | 🗸 | 🗸 |
| Clotrimazole | 🗸 | 🗸 | 🗸 |
| Clozapine | 🗸 | 🗸 | 🗸 |
| Cocaine | 🗸 | 🗸 | 🗸 |
| Colchicine | 🗸 | 🗸 | 🗸 |
| Cortisol | 🗸 | 🗸 | 🗸 |
| Csa | 🗸 | 🗸 | 🗸 |
| Cyclobenzaprine | 🗸 | 🗸 | 🗸 |
| Cyclophosphamide | 🗸 | 🗸 | 🗸 |
| Cyclosporin a | 🗸 | 🗸 | 🗸 |
| Cyclosporine | 🗸 | 🗸 | 🗸 |
| Cyproheptadine | 🗸 | 🗸 | 🗸 |
| D4t | 🗸 | 🗸 | 🗸 |
| Dacarbazine | 🗸 | 🗸 | 🗸 |
| Danazol | 🗸 | 🗸 | 🗸 |
| Dasatinib | 🗸 | 🗸 | 🗸 |
| Deferoxamine | 🗸 | 🗸 | 🗸 |
| Desipramine | 🗸 | 🗸 | 🗸 |
| Desvenlafaxine | 🗸 | 🗸 | 🗸 |
| Deucravacitinib | 🗸 |  |  |
| Dexfenfluramine hydrochloride | 🗸 | 🗸 | 🗸 |
| Dextroamphetamine | 🗸 | 🗸 | 🗸 |
| Dhea | 🗸 | 🗸 | 🗸 |
| Diclofenac | 🗸 |  |  |
| Dicyclomine | 🗸 | 🗸 | 🗸 |
| Dienestrol |  | 🗸 |  |
| Digoxin |  | 🗸 |  |
| Dihydroergocristine | 🗸 | 🗸 | 🗸 |
| Dihydroergotamine | 🗸 | 🗸 | 🗸 |
| Dilevalol hydrochloride | 🗸 | 🗸 | 🗸 |
| Dinoprost tromethamine | 🗸 | 🗸 | 🗸 |
| Dinoprostone | 🗸 | 🗸 | 🗸 |
| Dipivefrin | 🗸 | 🗸 | 🗸 |
| Dipivefrin hydrochloride | 🗸 | 🗸 | 🗸 |
| Dipyridamole | 🗸 | 🗸 | 🗸 |
| Disulfiram |  | 🗸 |  |
| Dobutamine | 🗸 | 🗸 | 🗸 |
| Dobutamine hydrochloride | 🗸 | 🗸 | 🗸 |
| Docetaxel | 🗸 | 🗸 | 🗸 |
| Dopamine | 🗸 | 🗸 | 🗸 |
| Dopamine hydrochloride | 🗸 | 🗸 | 🗸 |
| Doxepin | 🗸 | 🗸 | 🗸 |
| Doxorubicin | 🗸 | 🗸 | 🗸 |
| Dronedarone | 🗸 | 🗸 | 🗸 |
| Drotrecogin alfa | 🗸 | 🗸 | 🗸 |
| Droxidopa | 🗸 | 🗸 | 🗸 |
| Duloxetine | 🗸 | 🗸 | 🗸 |
| E-3810 | 🗸 |  | 🗸 |
| E-doxepin | 🗸 | 🗸 | 🗸 |
| Ecallantide |  | 🗸 | 🗸 |
| Efalizumab | 🗸 | 🗸 | 🗸 |
| Egcg | 🗸 | 🗸 | 🗸 |
| Enalapril |  | 🗸 |  |
| Enoximone | 🗸 | 🗸 | 🗸 |
| Entrectinib | 🗸 | 🗸 | 🗸 |
| Ephedrine hydrochloride | 🗸 | 🗸 | 🗸 |
| Ephedrine sulfate | 🗸 | 🗸 | 🗸 |
| Epinastine | 🗸 | 🗸 | 🗸 |
| Epinephrine | 🗸 | 🗸 | 🗸 |
| Epinephrine bitartrate | 🗸 | 🗸 | 🗸 |
| Epo | 🗸 | 🗸 | 🗸 |
| Epoprostenol | 🗸 | 🗸 | 🗸 |
| Epoprostenol sodium | 🗸 | 🗸 | 🗸 |
| Eptifibatide |  | 🗸 |  |
| Erbitux | 🗸 | 🗸 | 🗸 |
| Erdafitinib | 🗸 |  | 🗸 |
| Ergoloid mesylates | 🗸 | 🗸 | 🗸 |
| Erlotinib | 🗸 |  | 🗸 |
| Erythromycin | 🗸 | 🗸 | 🗸 |
| Esmolol | 🗸 | 🗸 | 🗸 |
| Esmolol hydrochloride | 🗸 | 🗸 | 🗸 |
| Estramustine | 🗸 | 🗸 | 🗸 |
| Etanercept | 🗸 | 🗸 | 🗸 |
| Etelcalcetide | 🗸 | 🗸 | 🗸 |
| Ethanol | 🗸 | 🗸 | 🗸 |
| Etoposide | 🗸 | 🗸 | 🗸 |
| Exemestane |  |  | 🗸 |
| Famciclovir | 🗸 | 🗸 | 🗸 |
| Famotidine | 🗸 |  | 🗸 |
| Fenoldopam | 🗸 | 🗸 | 🗸 |
| Fenoldopam mesylate | 🗸 | 🗸 | 🗸 |
| Fenoterol |  | 🗸 |  |
| Fenretinide | 🗸 | 🗸 | 🗸 |
| Fentanyl | 🗸 | 🗸 | 🗸 |
| Finasteride | 🗸 | 🗸 | 🗸 |
| Fk 506 | 🗸 | 🗸 | 🗸 |
| Flecainide | 🗸 | 🗸 | 🗸 |
| Fludarabine | 🗸 | 🗸 | 🗸 |
| Fluorouracil | 🗸 | 🗸 | 🗸 |
| Fluoxetine | 🗸 | 🗸 | 🗸 |
| Fluphenazine | 🗸 | 🗸 | 🗸 |
| Fluticasone propionate | 🗸 | 🗸 | 🗸 |
| Fluvoxamine | 🗸 |  | 🗸 |
| Folic acid | 🗸 | 🗸 | 🗸 |
| Formoterol |  | 🗸 |  |
| Formoterol fumarate |  | 🗸 |  |
| Foscarnet | 🗸 | 🗸 | 🗸 |
| Futibatinib | 🗸 |  | 🗸 |
| Gallium nitrate | 🗸 |  |  |
| Gemcitabine | 🗸 | 🗸 | 🗸 |
| Genistein | 🗸 | 🗸 | 🗸 |
| Glatiramer acetate | 🗸 |  |  |
| Glycopyrronium bromide |  | 🗸 |  |
| Gw642444 |  | 🗸 |  |
| Haloperidol | 🗸 | 🗸 | 🗸 |
| Hesperetin | 🗸 |  |  |
| Hexadecylphosphocholine | 🗸 | 🗸 | 🗸 |
| Htf 919 | 🗸 | 🗸 | 🗸 |
| Human c1-esterase inhibitor |  | 🗸 | 🗸 |
| Human chorionic gonadotropin | 🗸 | 🗸 | 🗸 |
| Hyaluronan | 🗸 | 🗸 | 🗸 |
| Hydrogen peroxide | 🗸 | 🗸 | 🗸 |
| Hydroquinone | 🗸 | 🗸 | 🗸 |
| Hydroxyamphetamine hydrobromide | 🗸 | 🗸 | 🗸 |
| Ibrutinib | 🗸 | 🗸 | 🗸 |
| Ibuprofen | 🗸 | 🗸 | 🗸 |
| Ici 182,780 | 🗸 |  | 🗸 |
| Idarubicin | 🗸 | 🗸 | 🗸 |
| Ifn alpha-2b | 🗸 |  |  |
| Iloperidone | 🗸 | 🗸 | 🗸 |
| Iloprost | 🗸 | 🗸 | 🗸 |
| Imatinib | 🗸 | 🗸 | 🗸 |
| Imipramine | 🗸 | 🗸 | 🗸 |
| Imiquimod | 🗸 |  |  |
| Incb54828 | 🗸 |  | 🗸 |
| Indacaterol | 🗸 | 🗸 | 🗸 |
| Indacaterol maleate |  | 🗸 |  |
| Indinavir | 🗸 | 🗸 | 🗸 |
| Indomethacin | 🗸 | 🗸 | 🗸 |
| Infigratinib | 🗸 |  | 🗸 |
| Infigratinib phosphate | 🗸 |  | 🗸 |
| Infliximab | 🗸 | 🗸 | 🗸 |
| Inositol | 🗸 |  |  |
| Insulin | 🗸 | 🗸 | 🗸 |
| Interferon alfa-2a | 🗸 |  |  |
| Interferon alfa-2a, recombinant | 🗸 |  |  |
| Interferon alfa-2b | 🗸 | 🗸 | 🗸 |
| Interferon alfa-2b, recombinant | 🗸 |  |  |
| Interferon alfa-n3 | 🗸 |  |  |
| Interferon alfacon-1 | 🗸 |  |  |
| Interferon alpha | 🗸 |  |  |
| Interferon alpha-2b | 🗸 | 🗸 | 🗸 |
| Interferon beta | 🗸 |  |  |
| Interferon beta-1a | 🗸 |  |  |
| Interferon beta-1b | 🗸 |  |  |
| Interleukin-11 |  | 🗸 |  |
| Irinotecan | 🗸 |  | 🗸 |
| Isoetharine | 🗸 | 🗸 | 🗸 |
| Isoetharine hydrochloride |  | 🗸 |  |
| Isoetharine mesylate |  | 🗸 |  |
| Isoproterenol | 🗸 | 🗸 | 🗸 |
| Isoproterenol hydrochloride | 🗸 | 🗸 | 🗸 |
| Isoproterenol sulfate | 🗸 | 🗸 | 🗸 |
| Isoxsuprine hydrochloride |  | 🗸 |  |
| Iudr | 🗸 |  |  |
| Kai-4169 | 🗸 | 🗸 | 🗸 |
| Ketotifen | 🗸 | 🗸 | 🗸 |
| Kx2-391 |  | 🗸 |  |
| Labetalol | 🗸 | 🗸 | 🗸 |
| Labetalol hydrochloride | 🗸 | 🗸 | 🗸 |
| Lactulose | 🗸 | 🗸 | 🗸 |
| Lanadelumab |  | 🗸 | 🗸 |
| Lansoprazole | 🗸 | 🗸 | 🗸 |
| Laropiprant | 🗸 | 🗸 | 🗸 |
| Leflunomide | 🗸 | 🗸 | 🗸 |
| Lenalidomide | 🗸 |  | 🗸 |
| Letrozole |  |  | 🗸 |
| Leucovorin | 🗸 | 🗸 | 🗸 |
| Levalbuterol |  | 🗸 |  |
| Levalbuterol hydrochloride |  | 🗸 |  |
| Levalbuterol tartrate |  | 🗸 |  |
| Levobetaxolol hydrochloride | 🗸 | 🗸 | 🗸 |
| Levobunolol | 🗸 | 🗸 | 🗸 |
| Levobunolol hydrochloride | 🗸 | 🗸 | 🗸 |
| Levodopa | 🗸 | 🗸 | 🗸 |
| Levosulpiride | 🗸 | 🗸 | 🗸 |
| Lifitegrast | 🗸 | 🗸 | 🗸 |
| Lithium | 🗸 | 🗸 | 🗸 |
| Lovastatin |  | 🗸 |  |
| Loxapine | 🗸 | 🗸 | 🗸 |
| Lurasidone | 🗸 | 🗸 | 🗸 |
| Luspatercept | 🗸 |  |  |
| Maprotiline | 🗸 | 🗸 | 🗸 |
| Masitinib | 🗸 | 🗸 | 🗸 |
| Mechlorethamine | 🗸 | 🗸 | 🗸 |
| Medroxyprogesterone | 🗸 | 🗸 | 🗸 |
| Medroxyprogesterone acetate | 🗸 | 🗸 | 🗸 |
| Melatonin | 🗸 | 🗸 | 🗸 |
| Melevodopa | 🗸 | 🗸 | 🗸 |
| Mephentermine sulfate | 🗸 | 🗸 | 🗸 |
| Metaproterenol sulfate |  | 🗸 |  |
| Methacholine |  | 🗸 |  |
| Methamphetamine |  | 🗸 |  |
| Methimazole | 🗸 | 🗸 | 🗸 |
| Methotrexate | 🗸 | 🗸 | 🗸 |
| Methylene blue | 🗸 | 🗸 | 🗸 |
| Methylergonovine | 🗸 | 🗸 | 🗸 |
| Methylergonovine maleate | 🗸 | 🗸 | 🗸 |
| Methylprednisolone | 🗸 | 🗸 | 🗸 |
| Methysergide | 🗸 | 🗸 | 🗸 |
| Metipranolol | 🗸 | 🗸 | 🗸 |
| Metipranolol hydrochloride | 🗸 | 🗸 | 🗸 |
| Metoclopramide hydrochloride | 🗸 | 🗸 | 🗸 |
| Metoprolol | 🗸 | 🗸 | 🗸 |
| Metoprolol fumarate | 🗸 | 🗸 | 🗸 |
| Metoprolol succinate | 🗸 | 🗸 | 🗸 |
| Metoprolol tartrate | 🗸 | 🗸 | 🗸 |
| Midazolam | 🗸 | 🗸 | 🗸 |
| Midostaurin | 🗸 |  | 🗸 |
| Mifepristone | 🗸 | 🗸 | 🗸 |
| Minaprine | 🗸 | 🗸 | 🗸 |
| Minaprine hydrochloride | 🗸 | 🗸 | 🗸 |
| Misoprostol | 🗸 | 🗸 | 🗸 |
| Mitoxantrone | 🗸 | 🗸 | 🗸 |
| Mmf | 🗸 | 🗸 | 🗸 |
| Molindone | 🗸 | 🗸 | 🗸 |
| Morphine | 🗸 | 🗸 | 🗸 |
| Mycophenolate mofetil | 🗸 | 🗸 | 🗸 |
| N-acetyl-l-cysteine | 🗸 | 🗸 | 🗸 |
| Nadolol | 🗸 | 🗸 | 🗸 |
| Naltrexone | 🗸 | 🗸 | 🗸 |
| Naproxen | 🗸 | 🗸 | 🗸 |
| Navelbine | 🗸 | 🗸 | 🗸 |
| Nebivolol | 🗸 | 🗸 | 🗸 |
| Nebivolol hydrochloride | 🗸 | 🗸 | 🗸 |
| Nelfinavir | 🗸 | 🗸 | 🗸 |
| Niacin | 🗸 | 🗸 | 🗸 |
| Nicardipine | 🗸 | 🗸 | 🗸 |
| Niclosamide | 🗸 | 🗸 | 🗸 |
| Nicotine | 🗸 | 🗸 | 🗸 |
| Nintedanib | 🗸 | 🗸 | 🗸 |
| Nintedanib esylate | 🗸 |  | 🗸 |
| Nitrogen mustard | 🗸 | 🗸 | 🗸 |
| Nivolumab | 🗸 |  | 🗸 |
| Nordihydroguaiaretic acid | 🗸 | 🗸 | 🗸 |
| Norepinephrine | 🗸 | 🗸 | 🗸 |
| Norepinephrine bitartrate | 🗸 | 🗸 | 🗸 |
| Nortriptyline | 🗸 | 🗸 | 🗸 |
| Ns-304 | 🗸 | 🗸 | 🗸 |
| Olanzapine | 🗸 | 🗸 | 🗸 |
| Olodaterol |  | 🗸 |  |
| Olodaterol hydrochloride |  | 🗸 |  |
| Omeprazole | 🗸 | 🗸 | 🗸 |
| Omidenepag | 🗸 | 🗸 | 🗸 |
| Omidenepag isopropyl | 🗸 | 🗸 | 🗸 |
| Oncovex gm-csf | 🗸 | 🗸 | 🗸 |
| Opc-34712 | 🗸 | 🗸 | 🗸 |
| Orciprenaline |  | 🗸 |  |
| Oxaliplatin | 🗸 | 🗸 | 🗸 |
| Oxprenolol | 🗸 | 🗸 | 🗸 |
| Oxprenolol hydrochloride | 🗸 | 🗸 | 🗸 |
| Oxtriphylline | 🗸 | 🗸 | 🗸 |
| Oxytocin | 🗸 | 🗸 | 🗸 |
| P-1101 | 🗸 |  |  |
| Paclitaxel | 🗸 | 🗸 | 🗸 |
| Palifermin | 🗸 |  | 🗸 |
| Paliperidone | 🗸 | 🗸 | 🗸 |
| Pamidronate | 🗸 | 🗸 | 🗸 |
| Panobinostat | 🗸 | 🗸 | 🗸 |
| Parathyroid hormone | 🗸 | 🗸 | 🗸 |
| Paroxetine | 🗸 | 🗸 | 🗸 |
| Pazopanib | 🗸 | 🗸 | 🗸 |
| Pazopanib hydrochloride | 🗸 |  | 🗸 |
| Peginterferon alfa-2a | 🗸 |  |  |
| Peginterferon alfa-2b | 🗸 |  |  |
| Peginterferon beta-1a | 🗸 |  |  |
| Pemigatinib | 🗸 |  | 🗸 |
| Penbutolol | 🗸 | 🗸 | 🗸 |
| Penbutolol sulfate | 🗸 | 🗸 | 🗸 |
| Pentamidine isethionate |  | 🗸 | 🗸 |
| Pentosan polysulfate | 🗸 |  | 🗸 |
| Pentoxifylline | 🗸 | 🗸 | 🗸 |
| Pergolide | 🗸 | 🗸 | 🗸 |
| Pergolide mesylate | 🗸 | 🗸 | 🗸 |
| Perphenazine | 🗸 | 🗸 | 🗸 |
| Phenobarbital | 🗸 | 🗸 | 🗸 |
| Phenylephrine | 🗸 | 🗸 | 🗸 |
| Phenylephrine hydrochloride |  | 🗸 |  |
| Pimozide | 🗸 | 🗸 | 🗸 |
| Pindolol | 🗸 | 🗸 | 🗸 |
| Pioglitazone | 🗸 |  |  |
| Pirbuterol |  | 🗸 |  |
| Pirbuterol acetate |  | 🗸 |  |
| Pirenzepine | 🗸 | 🗸 | 🗸 |
| Pirfenidone | 🗸 |  |  |
| Pirtobrutinib | 🗸 | 🗸 | 🗸 |
| Plegridy | 🗸 |  |  |
| Ponatinib | 🗸 | 🗸 | 🗸 |
| Practolol | 🗸 | 🗸 | 🗸 |
| Pravastatin | 🗸 | 🗸 | 🗸 |
| Prednisolone | 🗸 | 🗸 | 🗸 |
| Prednisone | 🗸 | 🗸 | 🗸 |
| Prilocaine hydrochloride |  | 🗸 |  |
| Procarbazine | 🗸 | 🗸 | 🗸 |
| Prochlorperazine | 🗸 | 🗸 | 🗸 |
| Promazine | 🗸 | 🗸 | 🗸 |
| Promethazine | 🗸 | 🗸 | 🗸 |
| Pronetalol | 🗸 | 🗸 | 🗸 |
| Propafenone | 🗸 | 🗸 | 🗸 |
| Propafenone hydrochloride | 🗸 | 🗸 | 🗸 |
| Propiomazine | 🗸 | 🗸 | 🗸 |
| Propranolol | 🗸 | 🗸 | 🗸 |
| Propranolol hydrochloride | 🗸 | 🗸 | 🗸 |
| Protokylol hydrochloride | 🗸 | 🗸 | 🗸 |
| Protriptyline | 🗸 | 🗸 | 🗸 |
| Prucalopride | 🗸 | 🗸 | 🗸 |
| Prucalopride succinate | 🗸 | 🗸 | 🗸 |
| Pseudoephedrine |  | 🗸 |  |
| Pyrogallol | 🗸 | 🗸 | 🗸 |
| Quercetin | 🗸 | 🗸 | 🗸 |
| Racepinephrine | 🗸 | 🗸 | 🗸 |
| Racepinephrine hydrochloride |  | 🗸 |  |
| Ramipril | 🗸 | 🗸 |  |
| Ranibizumab | 🗸 | 🗸 | 🗸 |
| Rapamycin | 🗸 | 🗸 | 🗸 |
| Recombinant gm-csf | 🗸 | 🗸 | 🗸 |
| Recombinant interferon alfa-2b | 🗸 |  |  |
| Recombinant interferon beta-1a | 🗸 | 🗸 | 🗸 |
| Regorafenib | 🗸 |  | 🗸 |
| Reproterol |  | 🗸 |  |
| Resveratrol | 🗸 | 🗸 | 🗸 |
| Retinoic acid | 🗸 | 🗸 | 🗸 |
| Retinol | 🗸 | 🗸 | 🗸 |
| Rhil-11 | 🗸 | 🗸 | 🗸 |
| Rhucin |  | 🗸 | 🗸 |
| Ribavirin | 🗸 | 🗸 | 🗸 |
| Rimiterol |  | 🗸 |  |
| Risperidone |  | 🗸 |  |
| Ritodrine |  | 🗸 |  |
| Ritodrine hydrochloride |  | 🗸 |  |
| Ritonavir | 🗸 | 🗸 | 🗸 |
| Rituximab | 🗸 |  |  |
| Ropeginterferon alfa-2b | 🗸 |  |  |
| Rotigotine | 🗸 | 🗸 | 🗸 |
| Salbutamol | 🗸 | 🗸 | 🗸 |
| Saline | 🗸 | 🗸 | 🗸 |
| Salmeterol | 🗸 | 🗸 | 🗸 |
| Salmeterol xinafoate |  | 🗸 |  |
| Salt |  | 🗸 |  |
| Saquinavir | 🗸 | 🗸 | 🗸 |
| Sar 1118.00 | 🗸 | 🗸 | 🗸 |
| Sargramostim | 🗸 | 🗸 | 🗸 |
| Selexipag | 🗸 | 🗸 | 🗸 |
| Selumetinib | 🗸 | 🗸 | 🗸 |
| Sertindole | 🗸 | 🗸 | 🗸 |
| Sertraline | 🗸 | 🗸 | 🗸 |
| Sirolimus | 🗸 |  | 🗸 |
| Sl-401 | 🗸 | 🗸 | 🗸 |
| Sodium salicylate | 🗸 | 🗸 | 🗸 |
| Sorafenib | 🗸 |  | 🗸 |
| Sotalol | 🗸 | 🗸 | 🗸 |
| Sotalol hydrochloride | 🗸 | 🗸 | 🗸 |
| Spironolactone |  | 🗸 |  |
| Sti 571 | 🗸 |  | 🗸 |
| Streptozocin | 🗸 | 🗸 | 🗸 |
| Streptozotocin | 🗸 | 🗸 | 🗸 |
| Stz | 🗸 | 🗸 | 🗸 |
| Sucralfate | 🗸 |  | 🗸 |
| Sunitinib | 🗸 | 🗸 | 🗸 |
| Sutimlimab |  | 🗸 | 🗸 |
| Tagraxofusp | 🗸 | 🗸 | 🗸 |
| Talc | 🗸 | 🗸 | 🗸 |
| Talimogene laherparepvec | 🗸 | 🗸 | 🗸 |
| Tamoxifen | 🗸 |  |  |
| Tegaserod | 🗸 | 🗸 | 🗸 |
| Tegaserod maleate | 🗸 | 🗸 | 🗸 |
| Temozolomide | 🗸 | 🗸 | 🗸 |
| Terbutaline |  | 🗸 |  |
| Terbutaline sulfate |  | 🗸 |  |
| Terfenadine | 🗸 | 🗸 | 🗸 |
| Testosterone | 🗸 |  |  |
| Testosterone undecanoate | 🗸 | 🗸 | 🗸 |
| Thalidomide | 🗸 | 🗸 | 🗸 |
| Theophylline | 🗸 | 🗸 | 🗸 |
| Theophylline sodium glycinate | 🗸 | 🗸 | 🗸 |
| Thiethylperazine | 🗸 | 🗸 | 🗸 |
| Thioridazine | 🗸 | 🗸 | 🗸 |
| Thrombin | 🗸 | 🗸 | 🗸 |
| Thyrotropin | 🗸 |  | 🗸 |
| Timolol | 🗸 | 🗸 | 🗸 |
| Timolol maleate | 🗸 | 🗸 | 🗸 |
| Tiotropium |  | 🗸 |  |
| Tirbanibulin |  | 🗸 |  |
| Tirofiban |  | 🗸 |  |
| Tirofiban hydrochloride |  | 🗸 |  |
| Tnt009 |  | 🗸 | 🗸 |
| Tocopherol acetate | 🗸 | 🗸 | 🗸 |
| Tofacitinib | 🗸 |  |  |
| Tofacitinib citrate | 🗸 |  |  |
| Toremifene | 🗸 |  |  |
| Tramadol |  | 🗸 |  |
| Trazodone | 🗸 | 🗸 | 🗸 |
| Treprostinil | 🗸 | 🗸 | 🗸 |
| Treprostinil diolamine | 🗸 | 🗸 | 🗸 |
| Triacetin | 🗸 | 🗸 | 🗸 |
| Triamcinolone | 🗸 |  | 🗸 |
| Trifluoperazine | 🗸 | 🗸 | 🗸 |
| Triflupromazine | 🗸 | 🗸 | 🗸 |
| Trimipramine | 🗸 | 🗸 | 🗸 |
| Troglitazone | 🗸 | 🗸 | 🗸 |
| Tropisetron | 🗸 | 🗸 | 🗸 |
| Tulobuterol |  | 🗸 |  |
| Upadacitinib | 🗸 |  |  |
| Ustekinumab | 🗸 | 🗸 | 🗸 |
| Vandetanib | 🗸 | 🗸 | 🗸 |
| Vargatef | 🗸 |  | 🗸 |
| Velcalcetide | 🗸 | 🗸 | 🗸 |
| Venlafaxine | 🗸 | 🗸 | 🗸 |
| Verapamil | 🗸 | 🗸 | 🗸 |
| Vidarabine | 🗸 | 🗸 | 🗸 |
| Vilanterol |  | 🗸 |  |
| Vilanterol trifenatate |  | 🗸 |  |
| Vinblastine | 🗸 | 🗸 | 🗸 |
| Vincristine | 🗸 |  | 🗸 |
| Vinorelbine | 🗸 | 🗸 | 🗸 |
| Vioxx | 🗸 | 🗸 | 🗸 |
| Vitamin D | 🗸 | 🗸 | 🗸 |
| Vitamin E | 🗸 |  |  |
| Vorinostat | 🗸 | 🗸 | 🗸 |
| Vortioxetine | 🗸 | 🗸 | 🗸 |
| Xamoterol | 🗸 | 🗸 | 🗸 |
| Yohimbine | 🗸 | 🗸 | 🗸 |
| Zanubrutinib | 🗸 | 🗸 | 🗸 |
| Zimeldine hydrochloride | 🗸 | 🗸 | 🗸 |
| Ziprasidone | 🗸 | 🗸 | 🗸 |
| Zolmitriptan | 🗸 | 🗸 | 🗸 |
| Zuclopenthixol | 🗸 | 🗸 | 🗸 |
